# Integrated immunogenomic analyses of high-grade serous ovarian cancer reveal vulnerability to combination immunotherapy

**DOI:** 10.1101/2024.01.09.24301038

**Authors:** Raphael Gronauer, Leonie Madersbacher, Pablo Monfort-Lanzas, Gabriel Floriani, Susanne Sprung, Alain Gustave Zeimet, Christian Marth, Heidelinde Fiegl, Hubert Hackl

## Abstract

**Background:** High-grade serous ovarian cancer (HGSOC) remains the most lethal gynecologic malignancy despite new therapeutic concepts, including poly-ADP-ribose polymerase inhibitors (PARPis) and antiangiogenic therapy. The efficacy of immunotherapies is modest, but clinical trials investigating the potential of combination immunotherapy with PARPis are underway. Homologous recombination repair deficiency (HRD) or BRCAness and the composition of the tumor microenvironment appear to play a critical role in determining the therapeutic response.

**Methods:** We conducted comprehensive immunogenomic analyses of HGSOC using data from several patient cohorts, including a new cohort from the Medical University of Innsbruck (MUI). Machine learning methods were used to develop a classification model for BRCAness from gene expression data. Integrated analysis of bulk and single-cell RNA sequencing data was used to delineate the tumor immune microenvironment and was validated by immunohistochemistry. The impact of PARPi and *BRCA1* mutations on the activation of immune-related pathways was studied *in vitro* using ovarian cancer cell lines, RNA sequencing, and immunofluorescence analysis.

**Results:** We identified a predictive 24-gene signature to determine BRCAness. Comprehensive analysis of the tumor microenvironment allowed us to identify patient samples with BRCAness and high immune infiltration. Further characterization of these samples revealed increased infiltration of immunosuppressive cells, including tumor-associated macrophages (TAMs) expressing *TREM2*, *C1QA,* and *LILRB4,* as identified by further analysis of single-cell RNA sequencing data and gene expression analysis of samples from patients receiving combination therapy with PARPi and anti-PD-1. PARPi activated the cGAS-STING signaling pathway and the downstream innate immune response in a similar manner to HGSOC patients with BRCAness status. We have developed a web application (https://ovrseq.icbi.at) and an associated R package OvRSeq, which allow for comprehensive characterization of ovarian cancer patient samples and assessment of a vulnerability score that enables stratification of patients to predict response to the mentioned combination immunotherapy.

**Conclusions:** Genomic instability in HGSOC affects the tumor immune environment, and TAMs play a crucial role in modulating the immune response. Based on various datasets, we have developed a diagnostic application that uses RNA sequencing data not only to comprehensively characterize HGSOC but also to predict vulnerability and response to combination immunotherapy.

## Background

Despite newer therapeutic concepts, ovarian cancer, particularly high-grade serous ovarian cancer, is still the deadliest gynecologic malignancy, with 13,270 expected deaths in 2023 in the U.S. [1]. While immunotherapy, such as immune checkpoint inhibition monotherapy (e.g., antibodies against PD-1 or PD-L1), has dramatically changed the therapeutic concepts of different cancer types, especially those with mismatch repair deficiency [2], the benefit for ovarian cancer patients with an objective response rate of approximately 10% was found to be rather modest [3–6]. However, poly-ADP-ribose polymerase inhibitors (PARPis) and antiangiogenic therapy have improved the survival outcomes of ovarian cancer patients beyond standard care, namely, debulking surgery and platinum-based therapy [7]. Furthermore, a number of clinical trials of combination therapies, including immune checkpoint blockade, are underway [8–12]. Whereas the recent primary analysis of the double-blind placebo-controlled ENGOT-Ov41/GEICO 69-O/ANITA phase III trial showed that the addition of the anti-PD-L1 antibody (atezolizumab) did not significantly improve the clinical outcome [12], early analysis of the MEDIOLA phase II study adding the PD-L1 inhibitor (durvalumab) and the angiogenesis inhibitor (bevacizumab) to a PARPi (olaparib) was promising, with an objective response rate >90% for a specific patient group with platinum-sensitive relapsed ovarian cancer harboring germline BRCA mutations [11].

PARP is involved in DNA damage and repair, binds to single-strand DNA breaks, and performs posttranslational modifications of histones and DNA-associated proteins by poly-ADP-ribosylation, also known as parylation. PARP inhibitors trap PARP and stall the replication fork, which can subsequently cause DSBs. PARP inhibition is synthetic lethal with deleterious *BRCA1* and *BRCA2* mutations because homologous recombination repair (HRR) cannot restore these double-strand breaks, introducing genome instability by nonhomologous end joining or leading to tumor cell death [13]. In high-grade serous ovarian cancer, approximately 14% harbor a germline and 6% a somatic mutation in the *BRCA1* or *BRCA2* gene, and approximately 50% are HRR deficient, indicating favorable PARPi therapy [14, 15]. Sequencing approaches such as targeted sequencing, whole exome sequencing, or whole-genome sequencing enable researchers to detect mutations in other genes involved in homologous recombination repair (HRR). However, the concept of HRR deficiency (HRD) or BRCAness goes beyond, as it encompasses instabilities and genomic scars, including large-scale transitions, loss of heterozygosity, telomeric allelic imbalance and specific mutational processes with uneven base substitution patterns (mutational signature 3). Several diagnostic assays from commercial providers for the detection of HRD have already been approved [16]. However, further efforts are undertaken to identify various biomarkers based on different modalities, such as gene expression or methylation, in the context of different cancer types [17–21]. Deleterious BRCA1 mutations and/or PARP inhibition can trigger an immune response at least in part through the cGAS-STING pathway [20-24], suggesting advantages for combined immunotherapies. However, biomarkers or phenotypes to predict the response to therapies, including PARPis and immune checkpoint blockers, are lacking.

In this study, we conducted comprehensive immunogenomic analyses of HGSOC using data from several patient cohorts, including a cohort from The Cancer Genome Atlas (TCGA-OV) (n=226), a new cohort from the Medical University of Innsbruck (MUI) (n=60), and data from the TOPACIO clinical trial in ovarian cancer patients treated with niraparib and pembrolizumab (n=22) (Fig. S1, Table S1, S2). Integrated gene expression analysis and machine learning on bulk and single-cell RNA sequencing data enabled the 1) development of a 24-gene expression classification model for BRCAness, 2) stratification of patient samples with BRCAness and high immune infiltration, whereby tumor-associated macrophages proved to be an important suppressive component, 3) identification of the activation of immune-related pathways such as the cGAS-STING or JAK-STAT pathway and downstream signaling by PARPi and *BRCA1* mutation (BRCAness), and 4) development of a diagnostic application from RNA sequencing data to comprehensively characterize HGSOC samples and predict vulnerability and response to combination immunotherapy.

## Methods

### Patient cohorts and datasets

The analysis workflow and used datasets from various cohorts are summarized in Fig. S1. Patient characteristics for the TCGA-OV cohort (n=226) and the new HGSOC cohort from the Medical University in Innsbruck (MUI) (n=60) are listed in Tables S1 and S2. RNA sequencing data and clinical data for the validation cohort (Medical University of Innsbruck; MUI) were deposited at https://doi.org/10.5281/zenodo.10251467. Controlled access data for whole exome sequencing and RNA sequencing data for the TCGA-OV cohort were obtained through dbGaP access permission (phs000178). Processed data (including methylation beta values) and clinical data were downloaded from Firebrowse (firebrowse.org), BROAD Institute). Additional clinical data were retrieved from the supplementary data of another resource [22]. Reads in bam format were converted into fastq files using samtools fastq [23]. Raw RNA sequencing data and clinical annotations for the ICON7 cohort were downloaded from the EGA archive (EGAS00001003487). Single-cell RNAseq data were downloaded from the Gene Expression Omnibus (GEO) (GSE180661) as an annotated count matrix (anndata-object) in h5ad-format. Data files from the TOPACIO clinical trial were retrieved from Synapse (https://doi.org/10.7303/syn21569629). RNA sequencing data for ovarian cancer cell lines were downloaded from the gene expression omnibus (GSE120792). Data from RNA sequencing analysis of OVCAR 3 and UWB1.289 cancer cell lines performed in this study were deposited in GEO (GSE237361).

### Cell line experiments

Two epithelial ovarian carcinoma cell lines, UWB1.289 harboring a deleterious *BRCA1* and OVCAR3 with intact *BRCA1,* were obtained from ATCC. OVCAR3 cells were grown in RPMI 1640 with 0.01 mg/ml bovine insulin and 20% FBS, whereas UWB1.289 cells were grown in a mixture of 48.5% MEGM Bullet Kit medium (Lonza) and 48.5% RPMI 1640 with 3% FBS. Viability assays were used to determine the IC50 for olaparib. Both cell lines were treated with olaparib or DMSO for 96 hours in four replicates. Treated and untreated UWB1.289 and OVCAR3 cells were stained with indirect immunofluorescent antibodies to detect γH2AX as an indicator of double-strand breaks. To determine activated STING signaling, double-stranded DNA and its presence in the cytosol, cGAS, STING, and phosphorylated STING were detected. The antibodies used are listed in Table S3.

### Immunohistochemistry analyses

Slices of 10 selected tumor blocks were subjected to immunohistochemistry analyses performed on the BenchMark ULTRA automated staining device (Ventana, Oro Valley, AZ/Roche, Vienna, Austria). The examined markers were CD163 for macrophages and CD8, PD-1, CD4, and FOXP3 for T cells. Furthermore, the markers γH2AX and STING were analyzed. All antibodies used are listed in Table S4.

### RNA sequencing analyses

RNA from cancer cell line samples was isolated from 2×10^6^ cells each using the RNeasy Mini Kit (Qiagen) according to the manufacturer’s protocols. RNA quantity and quality were assessed using NanoDrop™ 2000c and Bioanalyzer 2100 with Agilent 6000 Nano Kit, RNA integrity numbers (RIN) were between 9.3 and 9.8, and cDNA libraries were generated using the QuantSeq 3’ mRNA-Seq Library Prep Kit (Lexogen) according to the manufacturer’s instructions. Paired-end sequencing (150 bp) was performed on a NovaSeq 6000 sequencing device at GENEWIZ/Azenta. RNA isolation from 60 fresh frozen tumor samples from the HGSOC validation cohort from the Medical University of Innsbruck was conducted in a similar manner at the Department of Obstetrics and Gynecology, resulting in sufficient quality (RIN factors from 6.4 to 9.9), and sequencing was performed at Novogene (Cambridge, UK) for paired end sequencing (PE150) on an Illumina NovaSeq 6000 sequencing device using TrueSeq (Illumina) strand-specific total RNA libraries.

### RNA sequencing data analyses

Raw reads were quality checked using FastQC [24], and the results were summarized with MultiQC [25]. Reads were mapped to the human reference genome version hg38 (GRch38) using STAR (version 2.7.1) in 2pass mode [26]. Gene level expression quantification was performed with featureCounts (version 2.0.0) [27] using GENCODE annotations (v36). Raw counts were normalized using TPM (transcripts per million). RNA sequencing raw data from the HGSOC cohort from the Medical University of Innsbruck and the ICON7 cohort were analyzed in the same way. For raw sequencing data of the cell lines, single-end reads were processed by trimming adapter and low-quality sequences using BBDuk with the parameters specified by Lexogen. The trimmed reads were mapped to the human reference genome version hg38 (GRch38) using STAR (version 2.7.9a) in 2-pass mode. Gene level expression quantification was performed with featureCounts (version 2.0.0) and GENCODE annotations (v38).

### Whole exome sequencing analyses and variant calling

Raw exome sequencing reads in fastq format were quality checked using FastQC, and the results were summarized with MultiQC. Reads of paired tumor and normal samples were mapped against the human reference genome version hg38 (GRch38) using BWA [28]. After mapping the aligned bam files were sorted with samtools sort, mate coordinates and insert sizes were added with samtools fixmate, and duplicates were removed with samtools markdup. Finally, index files were generated using the SAMtools index. From exome sequencing data, small nucleotide variants (SNVs) consisting of single nucleotide variants and small indels were assessed. For germline variants, HaplotypeCaller was used to call variants with allele frequencies of 0.5 or 1.0 in the tumor samples as well as in the normal samples, respectively. To assess somatic variants in the tumor samples, four different variant callers, Mutect2 [29], SomaticSniper [30], Varscan2 [31], and Strelka2 [32], were used. If a variant was called by two of four variant callers and the variant allele frequency was ≥ 0.05 in the tumor sample and <0.05 in the normal sample, the variant passed filtering. Variants were annotated using VEP [33] with the ClinVar extension. Only pathogenic (class V) and likely pathogenic (class IV) variants were considered to affect the function of homologous recombination repair genes such as *BRCA1* or *BRCA2*. Tumor mutational burden was calculated based on the number of nonsynonymous single nucleotide variants per megabase for each tumor sample. Neoantigen prediction is based on previous efforts using analyses of a combination of exome and RNA sequencing data. HLA alleles (HLA type) were estimated using OptiType [34]. Variant calls, tumor and paired normal alignments, and aligned RNA sequencing reads were used together to compute mutational haplotypes using phasing. Based on the mutational haplotypes, peptides with lengths between 8-11 amino acids were generated and tested for the respective HLA alleles with NetMHCpan-4.0 [35], whereby %rank<2 was considered a weak binder and %rank<0.5 was considered a strong binder. Dissimilarity to the normal human proteome (hg38) was identified by the antigen.garnish package [36]. Neoantigen load was calculated for each tumor based on predicted weak and strong binding neoantigens – irrespective of their peptide length and taking all HLA alleles (type) into account – per megabase.

### Functional analysis of gene expression and the tumor immune environment

Differential gene expression analysis was conducted using the R package DESeq2 [37]. P values were adjusted for multiple testing based on the false discovery rate (FDR) according to the Benjamini‒Hochberg method. Genes with more than a twofold change at an FDR<0.1 and average expression across all samples (baseMean>10) were considered significantly differentially expressed. To identify functional annotation and affected biological processes, log2-fold change preranked gene set enrichment analyses (GSEA) [38] using hallmark and selected immune-related gene sets from MSigDB were performed. Overrepresentation analyses for biological processes (GO) and pathways (Reactome) were performed using the R package ClusterProfiler separately for significantly up- and downregulated genes [39]. ClueGO was used to build a network and group significantly overrepresented pathways, which are shared genes [40]. The STRING database [41] was used to identify an interaction network within the differentially expressed genes, and subnetworks were found by MCL clustering with inflation parameter=3. Footprint analyses of response genes of perturbed cancer signaling pathways were performed using PROGENy [42]. To assess tumor infiltration of immune cells, deconvolution methods, i.e., quanTIseq [43] using the immunedeconv R package [44] was applied to bulk RNA sequencing data from tumor samples. To characterize the immune-related processes, well-described immune signatures, such as T-cell inflammation, IFN gamma signature, cytolytic activity, cytotoxic T lymphocyte function, and T-cell exhaustion (Table S5), were analyzed. Based on log2(TPM+1) normalized expression data, single sample gene set enrichment using GSVA [45] was performed for signatures with more than 10 genes. For short signatures, including fewer than 10 genes, the average expression of the signature was calculated. The tumor-immune phenotype (infiltrated, excluded, desert) was determined based on a previously developed classification model based on 157 genes using digital pathology describing the presence and position of CD8+ T cells relative to the center or margin of the tumor [46]. A random forest model based on the expression data (TPM) of these 157 genes was used to characterize samples from the TCGA and the MUI cohorts. To classify ovarian cancer samples into immune reactive (IMR), proliferative (PRO), differentiated (DIF) and mesenchymal (MES) molecular subtypes, the consensusOV R package [47] was used. The immunophenoscore (IPS) and immunophenogram for all samples were determined as described previously [48].

### Determination of BRCAness

BRCAness was determined based on HRD scores [49], mutational signature 3 [50], mutations in homologous recombination repair pathway genes and methylation of promoter regions of *BRCA1* and *BRCA2*. All high-grade serous ovarian cancer (HGSOC) samples of the TCGA cohort for which paired tumor and normal exome sequencing and matched RNA sequencing data were available (n=226) were used. Samples were classified with a BRCAness phenotype when they had either a deleterious mutation in the homologous recombination pathway, an ovarian cancer-specific HRD score of ≥ 63 [51], a mutational signature 3 ratio > 0.25 or a methylation level beta value >0.7 of the BRCA1 or BRCA2 promoter. HRD scores were calculated as the unweighted sum of the three genomic scar values, loss of heterozygosity (LOH) [52], telomeric allelic imbalance (TAI) [53], and large-scale state transitions (LST) [54]. To compute the genomic scar values, scarHRD [55] was used on genome segmentation files generated with sequenza-utils [56]. The mutational signature 3 score was computed using MutationalPatterns [57]. The mutational signature 3 ratio was calculated as the ratio between mutational signature 3 supporting mutations and all detected mutations.

### BRCAness classification

First, genes expressed in ovarian cancer cells were identified using single-cell RNAseq data. Genes that are expressed in at least one ovarian cancer cell were considered expressed in cancer cells. Log_2_(TPM+1) normalized gene expression values of these genes in the TCGA dataset were then subjected to recursive feature elimination with three different machine learning models (random forest, AdaBoost and gradient boosting) to identify the 50 most important features for each model. Genes that were among the top 50 in at least two of the three models (24 genes) were then used subsequently to train a random forest classification model to discriminate between BRCAness and noBRCAness samples based on gene expression data. The performance of the classifier was evaluated by analysis of the receiver operating characteristic (ROC) curve with 10-fold cross-validation. The area under curve (AUC) was used as a performance measure. A cutoff for BRCAness (*P*>0.5266) was selected using the Youden index. Furthermore, the classifier was tested in the independent validation cohort (MUI) for 29 patients with HRD information based on SNP arrays and further validation using Myriad MyChoice CDx. Samplewise BRCA classification in single-cell RNA sequencing data from 29 patients was performed with an optimized cutoff (*P*>0.45) and based on the majority of classified tumor cells. The BRCAness classifier was further validated using additional somatic mutation and RNA sequencing data from the Clinical Proteomic Tumor Analysis Consortium ovarian cancer (CPTAC-OV) cohort [58]. SigMA scores representing mutational signature 3 were calculated using SigMA [59] as a surrogate for BRCAness.

### Single-cell RNA sequencing analysis

All analyses of single-cell data were performed in Python using scanpy [54] and scvi-tools [60]. Since the samples were sequenced separately for sorted CD45+ and CD45-cells, the raw read counts were integrated using scvi-tools with a batch effect correction. Counts were normalized to counts per million (CPM) and log2 transformed, adding a pseudocount of 1. Quality metrics were determined using scanpy and filtered for genes that are expressed in at least one cell. The dataset was filtered for samples from the primary tumor (adnexal tumor tissue). Principal component analysis and nearest neighbor analyses were calculated with default settings, and clustering was performed with the Leiden algorithm. Super cell types were annotated as previously defined. Subtypes of T-cell and myeloid cell clusters were assigned based on the expression of marker genes using published marker genes for different cell types and the PanglaoDB [61]. Differentially expressed genes between clusters were calculated using the Wilcoxon ranked sum test. For visualization, we used uniform manifold approximation and projection (UMAP) dimensional reduction. Gene expression between cell types was compared by heatmaps, violin plots, and bubble plots. To assess ligand‒receptor interactions between cancer cells and cells from the TME, CellPhoneDB [62] analysis was used.

### Gene expression analysis

Gene expression analysis in the TOPACIO cohort was performed on the NanoString platform. We used the nSolver software from NanoString (Seattle, US) to obtain normalized data. Differential expression analysis was performed using the R package limma [63], and genes with p<0.05 were considered differentially expressed.

### Vulnerability score and maps

Vulnerability maps consist of three variables: the vulnerability score, the BRCAness score and the cytolytic activity (CYT) to *C1QA* ratio. For the BRCAness score, the prediction probability from the random forest classifier was used. The CYT to *C1QA* ratio was calculated from the log_2_ (TPM+1) values of *GZMB*, *PRF1*, and *C1QA* (1).

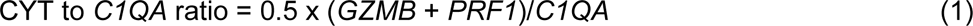

The CYT to *C1QA* ratio was transformed to values between 0 and 1 using a sigmoid function with softmax transformation and parameters derived from the TCGA cohort and termed C2C (2).

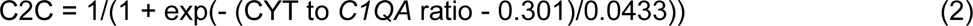

The vulnerability score was defined as the weighted sum of BRCAness probability and C2C (3), whereby the weights were identified using a logistic regression model on the CYT to *C1QA* ratio using log2 intensity expression values and SigMA status (mutational signature 3) data from the TOPACIO cohort and the treatment response as a binary dependent variable.

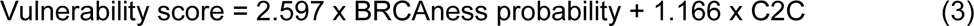

For visualization of the vulnerability map, a two-dimensional map was created with C2C as one coordinate, BRCA probability as the other coordinate, and the color-coded vulnerability score.

### Statistical analysis

Survival analyses were performed for both HGSOC cohorts (TCGA, MUI) for selected genes or immune parameters by dichotomization of patients based on the median or maximum log-rank statistics using the R package survival. For the TCGA cohort, overall survival and survival status were derived from a clinical data resource for TCGA [22] and for the cohort from Medical University Innsbruck from the clinical data as provided by the Department of Obstetrics and Gynecology. Univariate Cox regression was performed, and for parameters significantly associated with overall survival, multivariable Cox regression taking clinical parameters into account (age, FIGO stage, grade, residual tumor) was performed. To determine the association between continuous or binary variables, point biserial correlation analysis was used. For the correlation between binary variables, the Phi coefficient and chi-square test were used, and for the correlation between continuous variables, Pearson’s correlation coefficient was used. To compare parameters between two groups, the Wilcoxon rank-sum test was used. Where indicated, p values were adjusted for multiple testing based on the false discovery rate (FDR) according to the Benjamini‒Hochberg method. *P*<0.05 or FDR<0.1 were considered significant.

## Results

### A 24-gene signature predicts BRCAness in HGSOC patients

Because the response to platinum-based chemotherapies or therapy with PARP inhibitors in ovarian cancer is not limited to patients with tumors harboring *BRCA1* or *BRCA2* mutations, we expanded the group of patients by using a genomic characterization termed BRCAness, which has very much in common with homologous recombination repair deficiency (HRD) status [64]. BRCAness status includes mutations of genes in the homologous recombination DNA repair pathway (HRR), genomic scars, loss of heterozygosity, telomeric allelic imbalance, or large-scale transitions, mutational signature 3, or promoter methylation of the *BRCA1* or *BRCA2* gene. We assessed these parameters based on whole exome sequencing data and methylation data from the TCGA OV cohort (Fig. 1A). Very few patients harboring HRR mutations or BRCA1/2 promoter methylation fell below the combination of the HRD cutoff (HRD>63) and the MutSig3 ratio cutoff (0.25), indicating a reasonable selection of the cutoff values (Fig. 1B). To identify BRCAness solely based on gene expression data, we developed a machine learning classifier that can discriminate between BRCAness and non-BRCAness samples using bulk and single-cell RNA sequencing data (Fig. 1A). Recursive feature elimination based on multiple models resulted in a BRCAness gene expression signature with 24 genes, which was used to train a random forest model discriminating between BRCAness and noBRCAness. The receiver operating characteristics (ROC) with 10-fold cross-validation on the training dataset showed an area under the curve (AUC) of 0.91±0.04 (Fig. 1D). Furthermore, we demonstrated that in addition to classifying bulk RNAseq samples from the validation cohort (MUI) (Fig. 1E) with an accuracy of 0.79, an F1-score of 0.86, and a positive prediction value of 0.86, the classifier is also capable of classifying samples from scRNAseq data at the sample level (Fig. 1F) with an accuracy of 0.86, an F1 score of 0.87 and a positive prediction value of 0.87. There was also good agreement with a recently defined gene expression-based HRDness signature including 173 up- and 76 downregulated genes [65] using a single sample gene set enrichment [45, 66] derived score in the TCGA cohort as well as the MUI validation cohort with Spearman’s rank correlation of ρ=0.72 (*P*<0.001) and ρ=0.63 (*P*<0.001), respectively (Fig. S2, S3). Interestingly, six genes from our 24-gene signature to classify BRCAness (*CCDC90B*, *CRABP2*, *FZD4*, *GPAA1*, *PRCP*, *SNRP1*) were also among the upregulated and two genes (*RAD17*, *LTA4H*) among the downregulated genes. The 24-gene BRCAness signature was further validated in the CPTAC-OV cohort (n=71) by comparison to SigMA (mutational signature 3) with a Spearman’s rank correlation of ρ=0.43 (*P*<0.001) (Fig. S4).

**Fig. 1.**
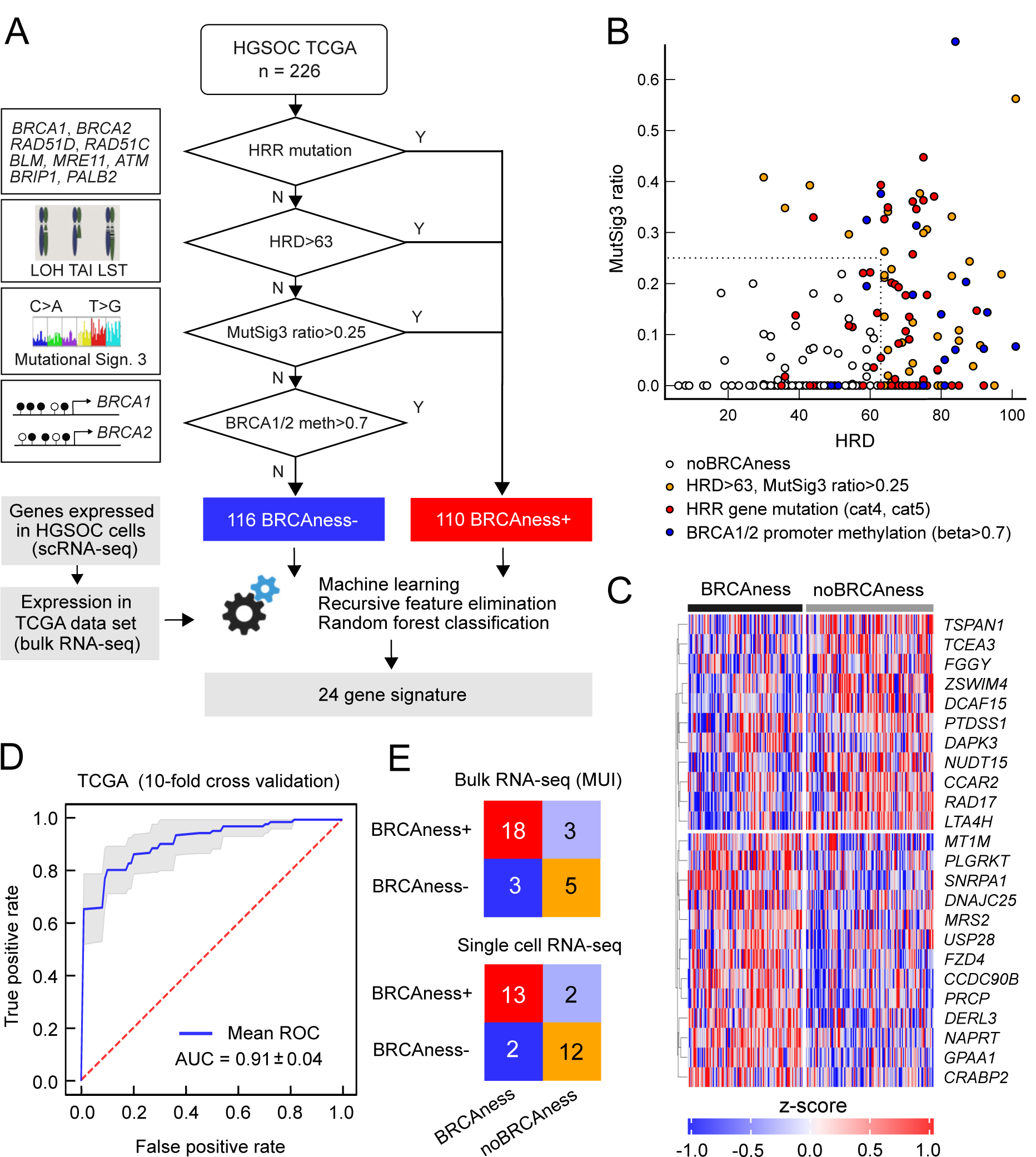
BRCAness classification based on the expression of 24 genes. **A** Decision tree of BRCAness determination in the TCGA-OV cohort and the development of a gene expression-based BRCAness classifier. **B** Different BRCAness parameters in the TCGA cohort compared between the HRD score and the mutation signature 3 ratio. Samples with mutated homologous recombination repair pathway genes are marked in red, BRCA1/2 promoter methylation in blue and samples with an HRD score > 63 and/or a signature 3 ratio > 0.25 but no mutation or BRCA1/2 promoter methylation are marked in yellow. Samples without BRCAness are marked in white. **C** Z scores of log_2_(TPM+1) normalized expression of the 24 genes of the BRCAness signature in the TCGA cohort as a heatmap clustered by BRCAness and non-BRCAness samples. **D** Mean ROC curve with 10-fold cross-validation of the classifier tested on the TCGA dataset. **E** Confusion matrices with correctly and incorrectly classified instances when the classifier was tested in independent test cohorts of single-cell RNA sequencing and bulk RNA sequencing data.

In summary, we developed a 24-gene-based BRCAness model validated in several single-cell and bulk RNAseq datasets with reasonable classification performance.

### Genome instability is associated with immune-related processes

To identify the relationship between genomic instability and the activation of the immune system, we performed correlation analyses between the BRCAness status and various immune-related signatures. BRCAness could be significantly positively associated with the enrichment of immune-related signatures, such as those for IFNG response (rho=0.38, p=0.004) and T-cell inflamed tumor microenvironment (rho=0.46, p=0.0014), even to a larger extent with high tumor mutational burden (p<0.001) and high neoantigen load (p<0.001) (Figure 2A, 2B). However, compared to other cancer types with defective DNA mismatch repair, such as melanoma or microsatellite instable colorectal cancer, the TMB or neoantigen load in ovarian cancer is rather low. Thus, this is more indicative of deficient homologous recombination repair. BRCAness was also associated with longer overall survival in the TCGA dataset (HR=0.50, 95%-CI 0.34-0.69; p<0.001 log rank test), indicating that those patients are more responsive to platinum-based chemotherapy (Fig. 2C). Although this status could be associated with higher CD8+ T-cell infiltration (estimated by deconvolution methods from RNA sequencing using quanTIseq) (HR=0.67, 95%-CI 0.47-0.93; p=0.019, log-rank test) (Fig. 2D), this could not completely explain the survival advantage. Nevertheless, analyses of signaling pathways by downstream target expression using PROGENy indicated for the TCGA cohort (n=226) as well as the MUI validation cohort (n=60) that immune-related pathways, including TNFa, NFkB, and JAK-STAT, were activated in the BRCAness samples (Fig. 2E, 2F). Using STRING analyses, we also identified a highly connected network including various chemokines and interleukins and their respective receptors (CCL7, CCL11, CXCL5, CXCL9, CXCL13, CCR2, CCR3, CCR4, CCR8, CXCR3, and IL6 (Fig. S11)), which were significantly more highly expressed in BRCAness tumors than in non-BRCAness tumors, indicating attraction and interaction with various immune cells.

**Fig 2.**
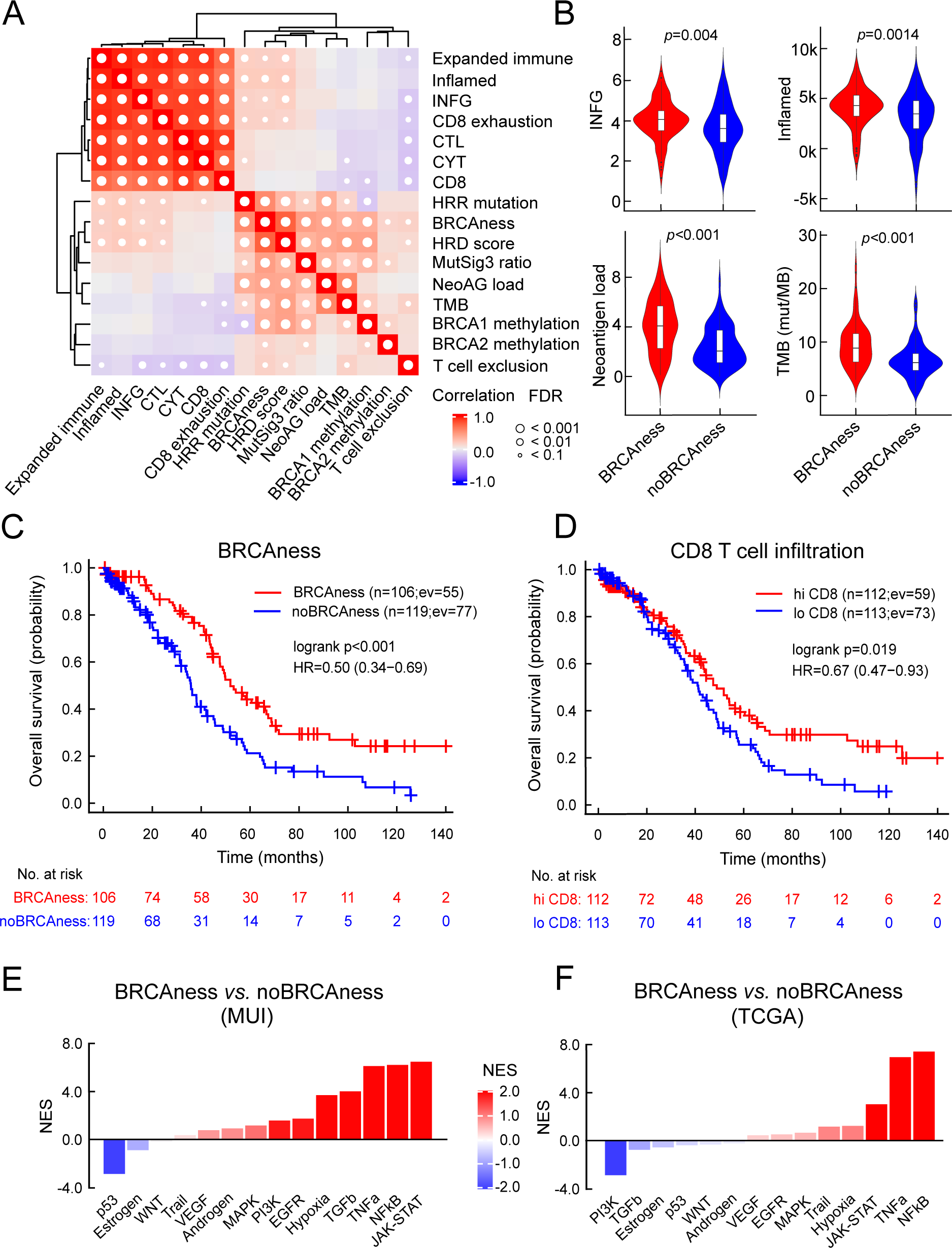
Association between BRCAness and immune parameters. **A** Results of correlation analysis of selected immune signatures and BRCAness parameters in the TCGA-HGSOC cohort (CYT, cytolytic activity; CTL, cytotoxic T lymphocytes; IFNG, interferon gamma signature; HRR mutations, mutations in the homologous recombination repair pathway; NeoAG load, neoantigen load; TMB, tumor mutational burden); white dots indicate significance (FDR<0.1). **B** Direct comparison of selected immune parameters between BRCAness and noBRCAness samples with significant differences, Wilcoxon rank-sum test (FDR<0.1). **C, D** Kaplan‒Meier curve for BRCAness and CD8 T-cell infiltration in the TCGA cohort. **E, F** Waterfall plot of normalized enrichment scores (NES) for the footprint analysis of immune-related pathways with PROGENy between BRCAness and non-BRCAness samples in the MUI (Medical University of Innsbruck) and TCGA cohorts.

We observed a significant association of BRCAness with longer overall survival and a less pronounced correlation with immune-related processes in HGSOC patients.

### PARP inhibition activates the cGAS-STING pathway *in vitro*

To study the effect of PARPis on immune activation, we performed *in vitro* analyses. As tumor models, an ovarian cancer cell line with a proficient BRCA1 gene (OVCAR3) and a cell line with a mutation in the BRCA1 gene (UWB1.289) were utilized. We performed RNA sequencing analyses to identify differentially expressed genes between olaparib (PARPi)-treated and control (DMSO)-treated cell lines. Significantly upregulated genes (Figure 3A, 3B, S18, S19, S20, S21, Data file 1) indicate activation of various processes (Fig. 3C, 3D, S22), including pattern recognition receptor activation, response to cytokine signaling, interferon alpha response (type I), NFkB pathway, and cGAS-STING signaling. To further validate the results at the protein level, we performed immunofluorescence analyses indicating effects on gH2AX by mutation in the BRCA1 gene and an even stronger effect by olaparib (PARPi) treatment (Fig. 3E). Similarly, we observed a different activation of cGAS and STING in the BRCA1-deficient versus the BRCA1-proficient cell model (Fig. 3F). Furthermore, using gene set enrichment analyses, a significant interferon alpha response was also observed in BRCAness samples of both the TCGA cohort and the MUI validation cohort (Fig. 3C).

**Fig 3.**
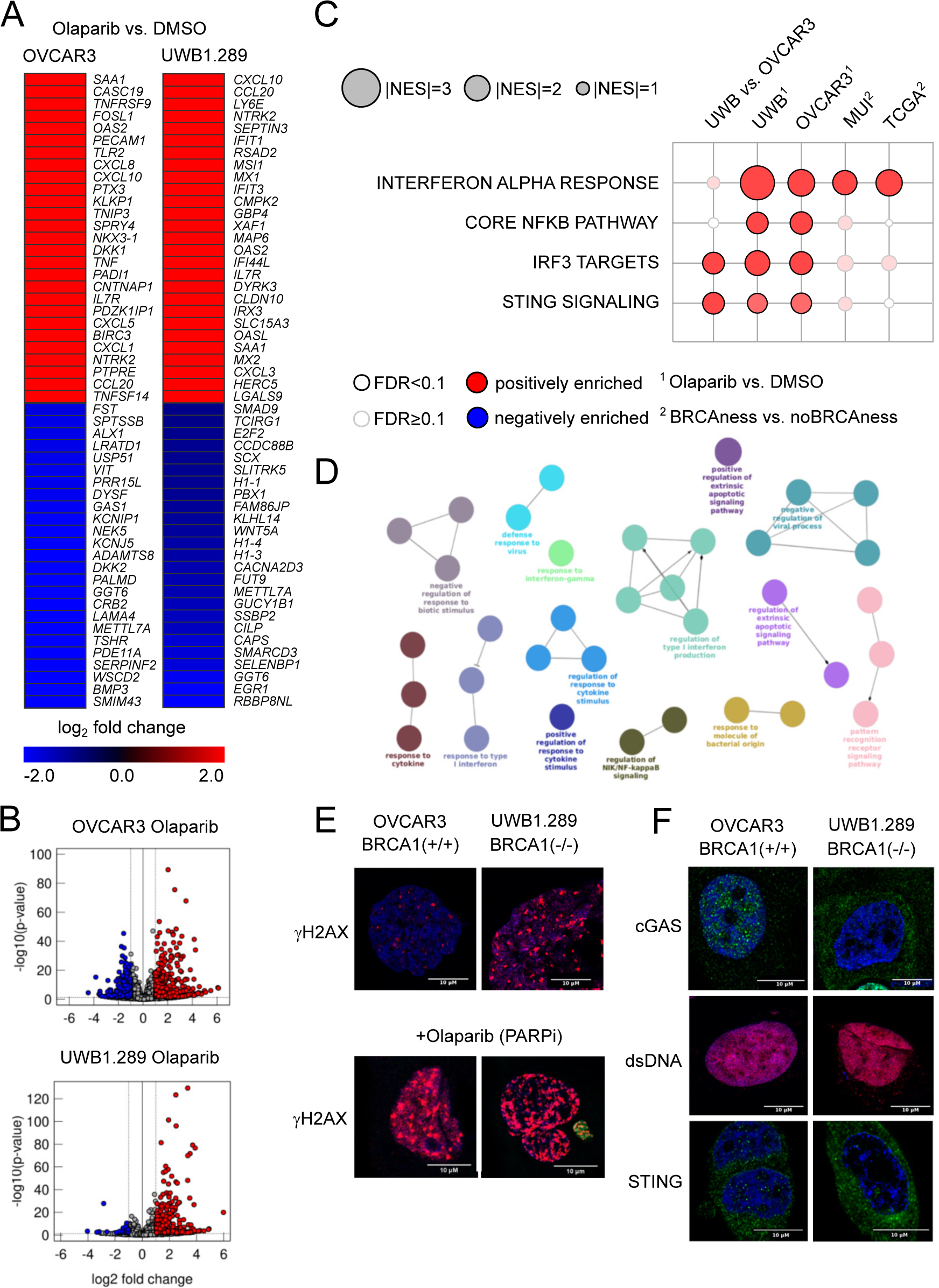
Results from cell line experiments with olaparib treatment **A** Top up- and downregulated genes for the cell lines OVCAR3 and UWB1.289 under olaparib treatment when compared to DMSO control. **B** General distribution of up- and downregulated genes after olaparib treatment compared to the DMSO control in both cell lines as volcano plots. Red indicates significantly upregulated genes (FDR<0.1, log_2_-fold change>1), and blue indicates significantly downregulated genes (FDR<0.1, log_2_-fold change<-1). **C** Normalized enrichment score of pathways associated with activation of the cGAS STING pathway in *BRCA1* mutated (cell lines) and BRCAness samples (cohorts) as well as olaparib-treated cell lines. **D** ClueGO network indicating overrepresented biological processes in the olaparib-treated UWB1.289 cell line. **E** Immunofluorescence staining of the DNA damage marker γH2AX in OVCAR3 and UWB1.289 cell lines with and without olaparib treatment. Comparing the different response to PARPi treatment between *BRCA1* mutation and wild type *BRCA1* **F** Immunofluorescence staining of cGAS, double stranded DNA (dsDNA) and STING in the OVCAR3 and UWB1.289 cell line comparing the difference between *BRCA1* mutation and wild type *BRCA1*.

In summary, we observed cGAS-STING activation by olaparib treatment *in vitro* and an interferon type I response as well as chemokine expression in HGSOC patient cohorts with BRCAness status.

### BRCAness and immune subtype stratifies HGSOC patients

We next focused on characterizing the presence of cytotoxic T lymphocytes and their spatial distribution in the tumor, following a recent approach in which digital pathology could be linked to gene expression [46]. With the reported list of 157 genes and using random forest analysis, we were able to divide the patients into a group with infiltrated, excluded, or desert tumor-immune phenotypes. Interestingly, the excluded phenotype was associated with upregulation of TGFβ and high expression of markers for cancer-associated fibroblasts, such as *FAP* or *PDPN*, which could form a physical barrier to prevent T-cell infiltration (Fig. 4A). Although various definitions of molecular subtypes based on gene expression or copy number aberrations have been described in the last decade, we are convinced that the immunoreactive molecular subtype (IMR) is the most meaningful to delineate immunoreactivity because many of the immunity genes, including cytotoxic effectors, factors involved in antigen processing and presentation, or immune checkpoints, are highly expressed in this condition (Fig. 4A, Table S6). The definition of molecular subtype also includes mesenchymal (MES), proliferative (PRO), and differentiated (DIF) molecular subtypes [47]. To identify patients most likely to benefit from the combination of PARP inhibitor therapy, where the BRCAness phenotype may be beneficial, with immune checkpoint inhibitor therapy, where the immune-related phenotype may be beneficial, we selected a group of patients with tumor BRCAness, an infiltrated tumor immune phenotype, and an immune-reactive molecular subtype termed BRCAness immune type (BRIT). When comparing the estimated immune cell infiltrates in these cancer samples with BRCAness cancers without immune type (noBRIT), we found that not only cytotoxic T lymphocytes such as CD8+ T cells were significantly more abundant (p<0.001) but also a number of suppressive immune cells (M2 macrophages (p<0.001), regulatory T cells (p<0.001), myeloid-derived suppressor cells; MDSCs (p<0.001)) (Fig. 4B). Furthermore, we did not observe a significant difference in overall survival between the groups (p=0.56, HR =0.81, 95% CI 0.42-1.60).

**Fig 4.**
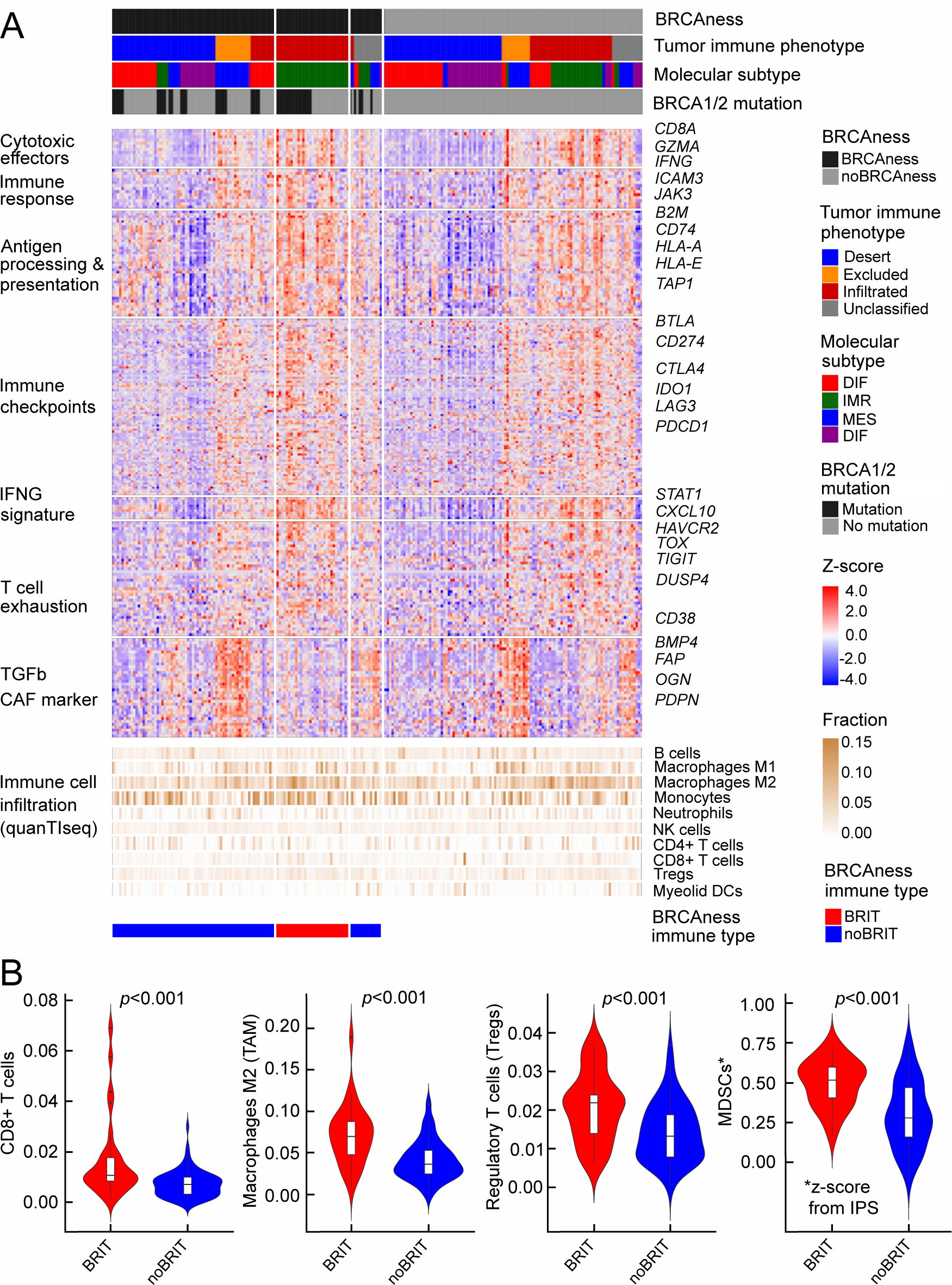
Profiles of immune parameters in the TCGA HGSOC cohort **A** Heatmap of z scores of log2(TPM+1) expression of immune-related genes and fraction of tumor infiltrating immune cells assessed with quanTIseq in all samples (n=226) from the TCGA-HGSOC cohort categorized by BRCAness, tumor-immune phenotype, molecular subtype and BRCA1/2 mutation. Furthermore, BRCAness samples are stratified into BRCAness immune type samples (BRIT), which show an immunoreactive molecular subtype and an infiltrated tumor-immune phenotype, and noBRIT samples, which only have BRCAness but do not fulfill the other two requirements. **B** Comparison of BRIT and noBRIT samples showing infiltration of CD8+ T cells, tumor-associated M2 macrophages (TAMs), regulatory T cells (Tregs) assessed with quanTIseq and myeloid-derived suppressor cells (MDSCs) assessed as the z score for MDSCs form the immunophenoscore (IPS).

These observations underscore the importance of the suppressive immune environment and suggest that suppressive immune cells may be an important factor, which is why ovarian cancer patients have a limited response to immunotherapy.

### Tumor-associated macrophages inform therapy response

As the power of deconvolution approaches from bulk RNA sequencing analyses shows some limitations, we took advantage of single-cell RNA sequencing analysis, allowing a more comprehensive characterization of the tumor environment and evaluation of the cell interplay. Analyses of more than 300 thousand cells of adnexal ovarian tumor tissue from 29 patients allowed a clear separation between major cell type populations by clustering and nonlinear projection (UMAP) (Figure 5A). In contrast to cell types from the tumor microenvironment, tumor cells showed a clear separation between BRCAness and noBRCness samples (Fig. 5A). Because cells from the suppressive environment have a major impact, we focused on the myeloid cell compartment and demonstrated that the majority of these cells were macrophages, and we identified subpopulations based on most dominant marker genes, including CD169 (SIGLEC1) macrophages, CX3CR1 macrophages, and MARCO macrophages (Fig. 5B). One described hallmark marker of tumor-associated macrophages (TAMs) is *TREM2,* which has been identified as an attractive target for cell depletion therapy and is being tested in an ongoing clinical trial [67]. Notably, the expression patterns of *TREM2* and BRCAness are very similar, showing high expression in all macrophage subtypes and, to a lesser extent, in monocytes (Fig. 5B). To search for further genes with similar expression patterns in myeloid subpopulations, we analyzed known tumor-associated macrophage and monocyte marker genes [68]. As indicated by this analysis, *C1QA* showed a similar but even more pronounced expression pattern than *TREM2* (Fig. 5B, 5C). *C1QA* was also recently described as a surrogate marker for the CD68+CD163+ macrophage subset [69].

**Fig 5.**
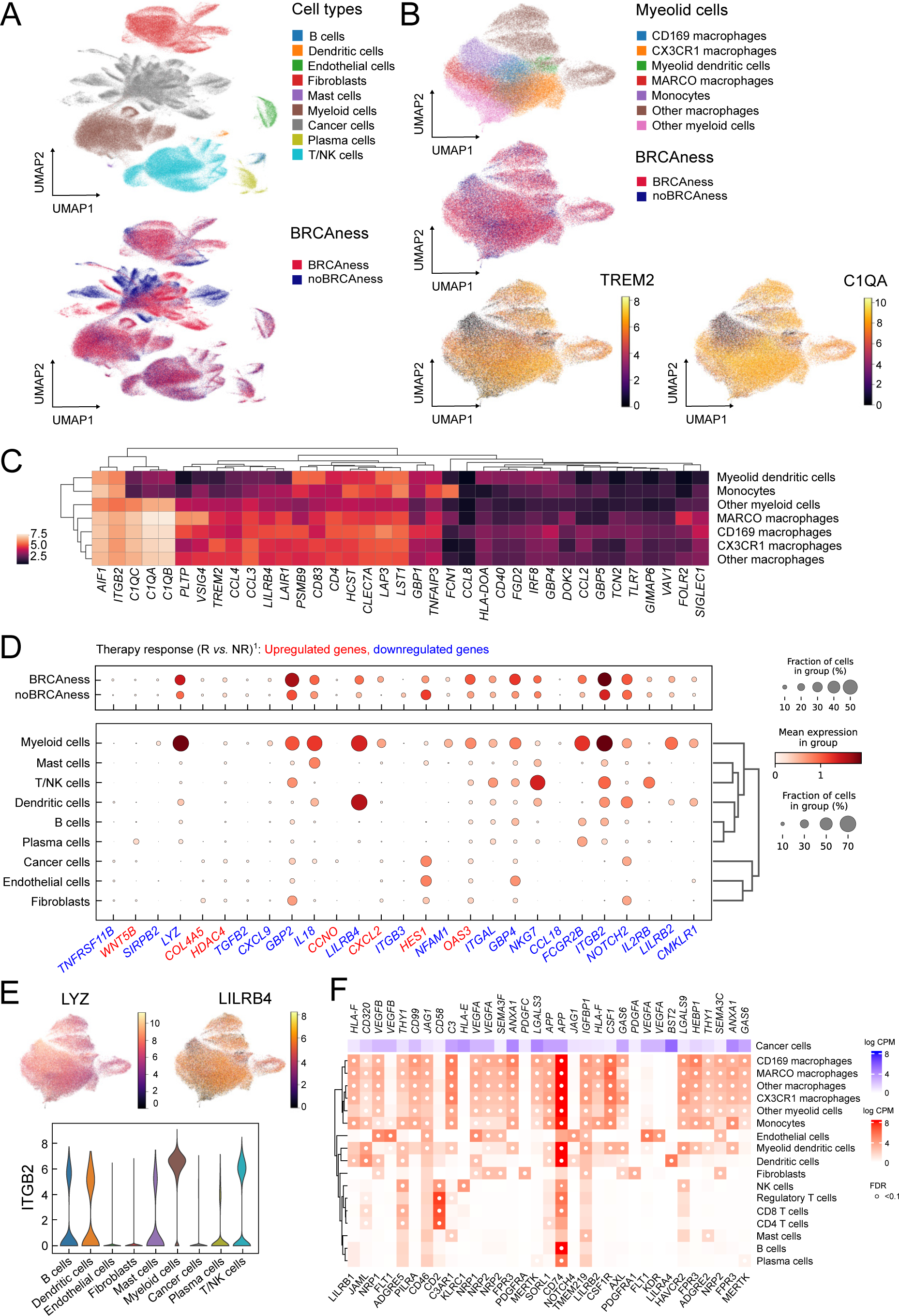
Single cell analysis of 29 ovarian cancer samples **A** UMAP showing the different cell types in of the ovarian cancer samples and which cells and cell types are associated with BRCAness samples. **B** UMAP plots of the myeloid cell compartment showing the association of macrophages with BRCAness cells and the expression of the macrophage marker gene *C1QA* and the TAM marker gene *TREM2* especially in cell clusters associated with BRCAness. **C** Heatmap of expression of macrophage associated marker genes in the different cell types in the myeloid cell compartment. **D** Association of genes identified as differentially expressed between responder and non-responder to PARPi-immune checkpoint inhibition combination therapy (niraparib and pembrolizumab) with different cell types and BRCAness. Genes depicted in blue are downregulated in responders while genes depicted in red are upregulated in responders. **E** UMAP visualization of *LYZ* and *LILRB4* genes highly expressed in the myeloid compartment, which are associated with BRCAness and non-responders. **F** Distribution of the expression of *ITGB2* in the different cell types, which is more highly expressed in non-responders and BRCAness.

To determine whether tumor-associated macrophages might also play a role in the response to combined cancer immunotherapy, we used expression data from a clinical trial (TOPACIO). We analyzed in which cell types genes with different expression in responders versus nonresponders were dominantly expressed. In fact, a number of downregulated genes in responders, such as *LYZ*, *LILRB4*, and *ITGB2,* were most highly expressed in myeloid cells (macrophages), *LILRB4* in dendritic cells, and integrin subunit beta 2 (*ITGB2*) in other cell types, such as T/NK cells (Fig. 5D, 5E). Interestingly, we identified various ligand‒receptor interactions with expressed ligands in tumor cells and respective receptors expressed in tumor-associated macrophage subsets using CellPhoneDB [62] (Fig. 5F).

The growth arrest-specific protein 6 (GAS6) – AXL tyrosine kinase (AXL) interaction, for example, which are both associated with poor outcome, have already been evaluated in clinical trials in ovarian cancer by inhibiting their interaction [70]. *LILRB1* and *LILRB2* expressed in macrophage subsets were found to interact with the nonclassical human leukocyte antigen *HLA-F* expressed in cancer cells. *VEGFA* and *VEGFB* expressed in tumor cells potentially interact with *NRP1*, and *FLT1 is* particularly expressed in endothelial cells. Blocking macrophage colony-stimulating factor *CSF1* and its receptor *CSF1R axis* and several drugs that target these factors have been under investigation [71].

These observations summarized together suggest that tumor-associated macrophages may not only play a role in immunotherapy alone but are also essential in informing about therapy response when combined with PARP inhibitors.

### Analyses of an independent cohort indicate vulnerability to combination immunotherapy

To validate the results, we performed RNA sequencing analyses of an HGSOC cohort of patients from Medical University Innsbruck (n=60). Stratification of these patients resulted in very similar expression patterns evident from a number of immune marker genes, which were highly expressed in the BRCAness immune type patient group (BRIT) (Fig. 6A). To further characterize immune infiltrates in different patient groups, we performed immunohistochemistry analyses on ten selected samples for various markers. BRIT tumor samples showed high γH2AX activity, STING activation, CD8+ T-cell infiltration, CD4+ T-cell infiltration, and strong CD163+ tumor-associated macrophage populations (Fig. 6B). These effects were even more pronounced in one sample with no detected BRCA1 or BRCA2 mutation, underscoring the importance and validity of predicted BRCAness. Another tumor sample with no BRCAness, a desert tumor-immune phenotype, and a differentiated molecular subtype was used as a negative control, and in fact, no activity for any of the tested markers was observed. To better address the potential for combination immunotherapy response, we again took advantage of data from the TOPACIO trial and, based on the clinical response, trained a logistic regression model and learned weights for three surrogate variables: MutSig3 as an indicator for BRCAness, average expression of *PRF1* and *GZMB* as indicators for cytolytic activity, and expression of *C1QA* as an indicator for tumor-associated suppressive macrophages. Based on the HGSOC samples from TCGA, we developed a two-dimensional vulnerability map, with the ratio of cytolytic activity and *C1QA* expression as one variable (C2C) and the BRCAness prediction probability as the other variable. The vulnerability score is indicated by color (Fig. 6C). When applied to the selected examples from the validation cohort of the Medical University of Innsbruck, these differed significantly for areas with high vulnerability scores (indicating response to combination immunotherapy) compared to the negative control with low vulnerability scores (Fig. 6C). Furthermore, we observed a significant difference in overall survival between patients with high and low vulnerability scores (p<0.001, HR = 0.47, 95% CI 0.33-0.66) in the TCGA HGSOC cohort, indicating a positive association of a high vulnerability score with longer overall survival. For patients in the validation cohort (MUI HGSOC), no significant difference in overall survival (p=0.368, HR = 0.78, 95% CI 0.45-1.34) could be revealed. To enable the characterization of newly diagnosed HGSOC samples based on RNA sequencing data, we developed an easy-to-use R package (OvRSeq), which allows us to not only estimate the parameters to determine the vulnerability score (and generate the vulnerability maps) but also comprehensively annotate the sample for BRCAness, tumor-immune phenotype, molecular subtype, estimate immune infiltrates, enrichment of immune-related signatures, and individual marker genes. This also includes other clinically relevant parameters, such as the angiogenesis score we previously defined, which might be useful for the prediction of anti-VEGF therapy [72]. The web application (https://ovarseq.icbi.at) allows the generation of summary information as a report of individual samples (Fig. S23).

**Fig 6.**
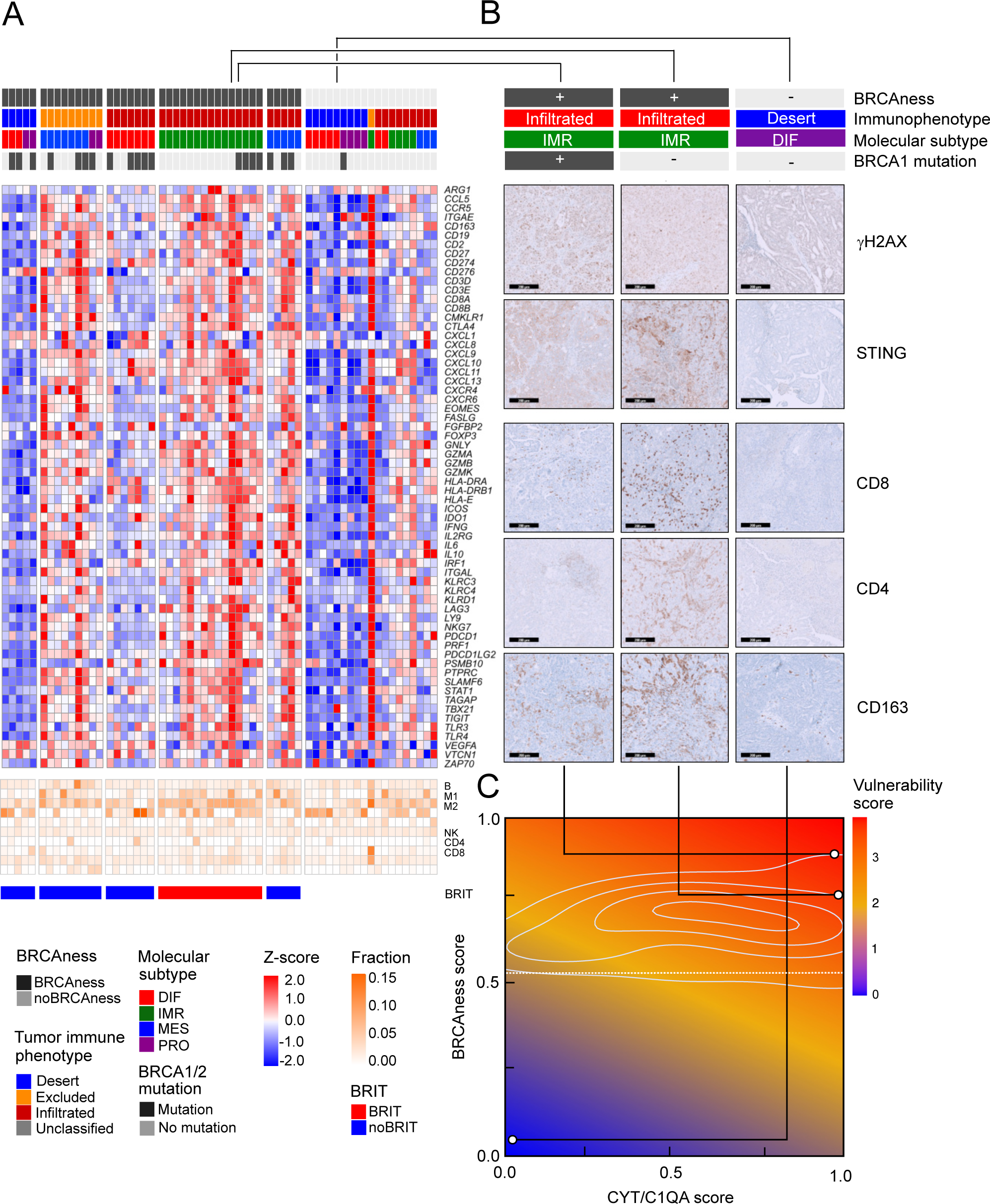
Expression profiles in the MUI cohort, immunohistochemistry validation, and vulnerability map **A** Heatmap of z-scores log_2_(TPM+1) expression of immune related genes and fraction of tumor infiltrating immune cells assessed with quanTIseq in all samples (n=60) from the MUI cohort categorized by BRCAness, tumor-immune phenotype, molecular subtype and BRCA1/2 mutation. **B** Immunohistochemistry images stained for CD8, CD4, CD163, FOXP3, γH2AX, and STING for three selected patients from the MUI cohort. Two BRIT samples one with a BRCA1 mutation and one without and one other sample without BRCAness, a deserted tumor-immune phenotype and a differentiated molecular subtype. **C** Vulnerability map showing the ratio between cytolytic activity CYT and *C1QA* (C2C) on the x-axis and the BRCAness score on the y-axis coloured by the vulnerability score. The three selected samples were mapped to the vulnerability map based on their CYT to *C1QA* ratio (C2C) and BRCAness score.

The developed application should ultimately be useful to identify vulnerabilities and support clinical therapy decisions for high-grade serous ovarian cancer patients.

## Discussion

Here, we described how genomic instability in high-grade serous ovarian cancer affects the tumor immune environment and the consequences and vulnerabilities of combination immunotherapy combining PARP inhibitors with immune checkpoint inhibitors such as anti-PD1 antibodies. A particular status in which patients respond well to PARP inhibitors and platinum-based chemotherapy is given when genes of the homologous recombination repair pathway such as *BRCA1* or *BRCA2* are mutated. Genomic scars are consequences of a homologous recombination repair deficiency and are used to define an HRD score, often measured by established commercial assays, which allows the assignment of a responsive status beyond *BRCA1* and *BRCA2* mutations. The applicability and associated cutoff values for different assays and cancer types are under discussion, as the HRD algorithm has been used in clinical studies including different cancer types, such as breast cancer and ovarian cancer [49,73,74]. Genomic scars are predictive but do not allow direct functional interpretation, whereas gene expression signatures could be an alternative in this regard. Very few approaches have associated gene expression with HRD status [18,19,65]. Whereas a sixty-gene signature [18] and a two-gene signature (*CXCL1*, *LY9*) [19] have focused on microarray data, a recent approach using RNA sequencing data identified a 249-gene signature to predict HRD [65]. We observed a number of overlaps with our 24-gene BRCA signature and a high concordance of signature scores in our training (TCGA) and validation (MUI) cohorts, indicating the reliability of our approach. This was underscored by comparison with mutational signature 3 (SigMA) in an independent cohort (Fig S3). The performance of the BRCAness classifier is reasonable, with AUC=0.91 (10-fold cross-validation) and positive predictive value for validation on both bulk RNA sequencing in the validation cohort (MUI) (PPV=0.86) and samplewise single-cell RNA sequencing data (PPV=0.87).

BRCAness is associated with a longer overall survival since all patients usually receive platinum-based chemotherapy such as carboplatin-paclitaxel combination. Although this association was previously reported and therefore expected, it highlights the performance of the classifier to BRCA status. There is evidence that BRCA1/2-mutated tumors exhibit significantly increased CD8+ TILs [75], although in breast cancer, differential modulation between BRCA1 and BRCA2 mutations in the tumor immune microenvironment has been found [76]. The association between BRCAness and several immune-regulated signatures was significant but not very pronounced. We found evidence that several signaling pathways and processes known to modulate the immune system are activated by BRCA1 mutations or a BRCAness-related phenotype, such as JAK-STAT signaling or an interferon type I response, which are activated by free double-stranded DNA in the cytoplasm of tumor cells via the cGAS-STING pathway and affect dendritic cells [77–79]. By expression and immunofluorescence analyses of ovarian cancer cell lines and by treatment with PARPi, we demonstrated that this axis is actually activated. Notably, the STAT3 pathway, which is activated by PARP inhibition, may, however, mediate treatment resistance by promoting the polarization of protumor TAMs, which could be overcome by STING agonism [80]. STING, CSF1R, SREBP-1, and VEGFA might also be targets to overcome resistance to PARPi-immunotherapy combinations [81]. The upregulation of many chemokines and chemokine receptors (Fig. S10, S11) indicates that BRCAness tumors are actively involved in immune cell attraction and interaction. For example, CCL5 produced by tumor cells or CXCL9 and CXCL10 also expressed by tumor-resident myeloid cells determine effector T-cell recruitment to the tumor microenvironment [82, 83]. We detected significant upregulation of CCL5 and CXCL10 by PARP inhibition, which was also identified as a downstream target of STING [77]. Another interesting chemokine that is strongly upregulated in cancer cell lines, particularly by olaparib treatment, is CCL20. CCL20 could be associated with cancer metastasis and progression by interacting with its cognate receptor CCR6 in an ovarian cancer mouse model. However, the higher expression in the myeloid cell compartment, as evident from single-cell analyses (Fig. S16), overlies the intrinsic tumor effect. One of our basic hypotheses was that samples with BRCAness respond better to PARPi therapy and that hot tumors with an activated immune milieu respond better to immune checkpoint inhibition, as has been shown, for example, in melanoma for the activated IFNG pathway [84]. However, when we compared the BRCAness immune type (BRIT) with other samples, we observed by using deconvolution approaches that suppressive cell types such as M2 macrophages, MDSCs, and Tregs were more abundant. In particular, tumor-associated macrophages (TAMs) could be a major factor together with low mutational burden, abnormal neovascularization, altered metabolism, and failure to reverse T-cell exhaustion for the limited immunotherapy response in ovarian cancer [85]. By using single-cell RNA sequencing data analyses in adnexal cancer tissue from 29 patients, we demonstrated that myeloid cells are the most abundant immune cells, and the majority were characterized as tumor-associated macrophages. We could identify various subtypes of these TAMs, and based on known macrophage polarization marker genes (Fig. S14), we observed a bias toward alternative (M2-like) macrophages compared to classical (M1-like) macrophages, although this classification is limited and may be better described as a continuum of different stages than isolated cell types. Nevertheless, a majority of these TAMs are suppressive, as indicated by *TREM2* expression. TREM2 is a promising therapeutic target for TAM depletion [71]. Inhibition of TREM2 has been shown to improve the anti-PD1 response in various mouse models and is currently being investigated in a clinical trial [67, 86]. Another recent study underscored the role of TAMs and demonstrated that specifically, the Siglec-9-positive TAM subset is associated with an immune-suppressive phenotype and adverse prognosis in HGSOC patients [87].

Interestingly, a previous work using cyclic immunofluorescence highlighted the role of exhausted T cells in the response to niraparib/pembrolizumab. In responders, particularly in extreme responders, frequent proximity between exhausted T cells and PD-L1+ (CD163+, IBA1+, CD11b+) TAMs was observed [9]. Noticeably, based on the selected marker expression, we observed an overlap with the CD169/SIGLEC-1 TAM cluster from single-cell RNA sequencing data analyses (Fig. S9). In addition, in patients who responded to this combination therapy, we identified a number of downregulated genes that were also highly expressed in TAMs, such as *LYZ*, *LILRB4*, and *ITGB2*. Whereas lysozyme (LYZ) is an antimicrobial ligand and is involved in central macrophage function and is therefore nonspecifically and highly expressed, *LILRB4* is an immune checkpoint on myeloid cells, indicating a more regulatory role. High expression of the integrin *ITGB2* was previously shown to be associated with poor survival outcome [88], underscoring that high expression in TAMs is crucial. In contrast, *ITGB2* is also associated with CD8+ T cells, as it encodes the beta chain of the LFA-1 protein, which has been shown to be essential in the assembly of the immune synapse or to influence lymphocyte extravasation and T-cell recruitment to the tumor and is regulated by GDF-15 [89]. This association is also underlined by a positive significant correlation of *ITGB2* expression with M2 macrophages and, to a lesser extent, with estimated CD8+ T-cell infiltration in the TCGA cohort (Fig. S11).

Because stratification of patients based on gene expression in our validation cohort was very similar to the analysis on the TCGA cohort, we set out to adapt our hypothesis andalso include elements of the suppressive environment. Already, it was shown that regulatory T cells (Tregs) are an important component of the suppressive milieu and are associated with unfavorable survival outcomes in ovarian cancer [90, 91]. We performed immunohistochemistry analyses using FOXP3 and CD163 antibodies in the validation cohort and found very pronounced macrophage infiltration (CD163) but hardly Treg infiltration (FOXP3) into the tumor site in some samples. The results of the single-cell RNA sequencing analyses and the fact that various TAM marker genes were associated with poorer overall survival (Fig. S13) also suggest that TAMs play a more dominant role in ovarian cancer.

While infiltration of various cell types from the adaptive immune system [92] and other markers, such as tumor mutational burden (TMB) [93] or IFNG signature [84], have been associated with good prognosis and immunotherapy response in various cancer types, the suppressive immune environment with tumor-supportive CD68+CD163+ macrophages is becoming more important [69]. Accordingly, a signature of the immune activation ratio of CD8A/C1QA has been found to be prognostic and predictive for immunotherapy response [69]. Based on previous analyses [44, 69], we considered the mean PRF1 and GZMB expression as a proxy for cytolytic activity as predominantly exerted by cytotoxic T lymphocytes. The specific expression pattern of C1QA on TAMs was comparable to that of TREM2 but at a much higher level. Therefore, we also used the member of the complement system C1QA as a surrogate for TAMs and the suppressive tumor immune environment and finally built a ratio of cytolytic activity (CYT) to the expression of C1QA (C2C), indicating the pro- and antitumoral balance of the immune environment. Finally, to build a predictive algorithm for combination therapy response, we included both C2C on the one hand and BRCAness on the other hand into one model. Since HRD measured with companion diagnostic tests is not able to predict all PARPi responders, as shown in several clinical trials, and since PARPi treatment can activate a number of immune-related pathways even in situations with proficient HRR, which is also underlined by our *in vitro* analyses, this model is considered to be relevant for combination immunotherapy.

Our studies have some limitations in that the training and validation patient cohorts were retrospective studies, and RNA sequencing was performed at a later time point. Additionally, only a limited number of patients who received combination therapy could be included; therefore, the conclusion about the predictive power for the treatment is limited and requires further validation in larger cohorts. One component that was not considered in this study is malignant ascites, which has been shown to contain various cell types, such as macrophages, many soluble factors and cytokines, that influence the protumorigenic phenotype and promote metastatic spread of HGSOC through transcoelomic dissemination [94]. Although the application is not a clinically approved software for the purpose of therapy and diagnosis, the very easy-to-use application (https://ovrseq.icbi.at) and the respective R package OvRSeq allow based on RNA sequencing to gain comprehensive information about the phenotype of a tumor sample, support clinical decisions, and stimulate further research.

## Conclusions

Our approach using RNA sequencing data to comprehensively characterize both genome instability and the tumor immune environment enabled us to stratify HGSOC patients. However, further analyses indicate that suppressive tumor-associated macrophages in the tumor immune microenvironment may play an essential role in understanding why immunotherapy shows only a modest response in ovarian cancer, and in a similar fashion, this applies to combination immunotherapy, including PARP inhibitors and immune checkpoint blockers. Based on various datasets, we have developed a methodology and corresponding diagnostic application that uses RNA sequencing data not only to comprehensively characterize newly diagnosed HGSOC patients but also to provide information on vulnerability to combination immunotherapy that may inform whether the patient will respond or not.

## Abbreviations

AUC: Area under curve
BRCA: Breast cancer DNA repair-associated genes
BRIT: BRCAness immune type
CTL: Cytotoxic T lymphocytes
CYT: Cytolytic activity
C1QA: Complement C1q A chain
C2C: Transformed CYT to *C1QA* ratio
DIF: Differentiated molecular subtype
EOC: Epithelial ovarian cancer
FDR: False discovery rate
GSEA: Gene set enrichment analysis
GSVA: Gene set variation analysis
HGSOC: High-grade serous ovarian cancer
HR: Hazard ratio
HRD: Homologous recombination repair deficiency
HRR: Homologous recombination repair
IMR: Immune reactive molecular subtype
IPS: Immunophenoscore
LOH: Loss of Heterogeneity
LST: Large-scale transitions
MDSC: Myeolid-derived suppressor cell
MES: Mesenchymal molecular subtype
MutSig3: Mutational signature 3
NES: Normalized enrichment score
PARP: Poly (ADP-Ribose) Polymerase
PARPi: PARP inhibitor (olaparib, niraparib
PCA: Principal component analysis
PD-1: Programmed cell death 1 (*PDCD1*)
PRO: Proliferative molecular subtype
PROGENy: Pathway RespOnsive GENes for activity inference
ROC: Receiver operating characteristics
TAI: Telomeric allelic imbalance
TAM: Tumor-associated macrophages
TCGA: The Cancer Genome Atlas
TPM: Transcript per millions
Tregs: Regulatory CD4+ T cells
UMAP: Uniform manifold approximation and projection

## Author contributions

RG conducted all computational analyses, and LM performed all *in vitro* analyses. PML developed the R package and application. GF performed antigen prediction and deconvolution analyses. SS performed immunohistochemistry analyses. AGZ served as a clinical consultant. CM headed the pilot study from the Medical University in Innsbruck and supervised all clinical aspects. HF coordinated tumor samples from the biobank and their molecular analysis. HH conceived the study, supervised all analyses, and together with RG wrote the paper. All the authors have read and approved the final manuscript.

## Ethics approval and consent to participate

For the pilot study (validation cohort from the Medical University of Innsbruck), written informed consent was needed for all patients before enrollment. The study was reviewed and approved by the Ethics Committee of the Medical University of Innsbruck (reference number: 1189/2019) and conducted in accordance with the Declaration of Helsinki.

## Availability of data and materials

RNA sequencing data from *in vitro* experiments are available via the Gene Expression Omnibus (GEO) (GSE237361). RNA sequencing data and patient information of the validation cohort (MUI) are available at Zenodo https://doi.org/10.5281/zenodo.10251467. The R package OvRSeq is available from GitHub (https://github.com/icbi-lab/OvRSeq) under MIT license and the corresponding web application (https://ovrseq.icbi.at). The analysis scripts used in this manuscript are available at GitHub (https://github.com/icbi-lab/hgsoc).

## Data Availability

RNA sequencing data from in vitro experiments are available via the Gene Expression
Omnibus (GEO) (GSE237361). RNA sequencing data and patient information of the validation
cohort (MUI) are available at Zenodo https://doi.org/10.5281/zenodo.10251467. The R
package OvRSeq is available from GitHub (https://github.com/icbi-lab/OvRSeq) under MIT
license and the corresponding web application (https://ovrseq.icbi.at). The analysis scripts
used in this manuscript are available at GitHub (https://github.com/icbi-lab/hgsoc).

