## Supplementary material for "Integrated immunogenomic analyses of high-grade serous ovarian cancer reveal vulnerability to combination immunotherapy"

### Supplementary Tables

**Table S1:** Patient characteristics of the high grade serous ovarian cancer (HGSOC) subset of the TCGA-OV cohort.

| Parameter | Range / Number of patients (n=226) (%) |
|---|---|
| Age | 34-87 |
| FIGO Stage | |
| 1 and 2 | 14 (6.2) |
| 3 | 173 (76.5) |
| 4 | 37 (16.4) |
| na | 2 (0.9) |
| BRCAness | 116 (51.3) |
| noBRCAness | 110 (48.7) |
| HRD-scores | 6-101 |
| >63 | 89 (39.4) |
| <=63 | 137 (60.6) |
| Patients with HRR mutation | 54 (23.9) |
| BRCA1 mutations (germline/somatic) | 25/3 (11.1/1.3) |
| BRCA2 mutations (germline/somatic) | 14/1 (6.2/0.4) |
| other HRR mutations | 13 (5.8) |
| Tumor immune phenotype | |
| Infiltrated | 79 (35.0) |
| Excluded | 27 (11.9) |
| Deserted | 97 (42.9) |
| Unclassified | 26 (11.5) |
| Molecular subtype | |
| Immunoreactive | 66 (29.2) |
| Differentiated | 69 (30.5) |
| Proliferative | 48 (21.2) |
| Mesenchymal | 43 (19.0) |

**Table S2:** Patient characteristics of the HGSOC cohort from the Medical University of Innsbruck (MUI).

| Parameter | Range / Number of patients (n=60) (%) |
|---|---|
| Age | 32-83 |
| FIGO Stage | |
| 3 | 48 (80.0) |
| 4 | 12 (20.0) |
| BRCAness | 41 (68.3) |
| noBRCAness | 19 (31.7) |
| BRCA1 mutations | 16 (26.7) |
| BRCA2 mutations | 4 (6.7) |
| Tumor immune phenotype | |
| Infiltrated | 36 (60.0) |
| Excluded | 10 (16.7) |
| Deserted | 14 (23.3) |
| Molecular subtype | |
| Immunoreactive | 20 (33.3) |
| Differentiated | 17 (28.3) |
| Proliferative | 8 (13.3) |
| Mesenchymal | 15 (25.0) |

**Table S3:** Antibodies used for immunofluorescence staining of ovarian cancer cells from the UWB1.289 and OVCAR3 cell lines.

| **Antibody** | **catalogue number** | **manufacturer** |
|---|---|---|
| ProLong® Gold Antifade Reagent with DAPI | #8961 | Cell Signaling Technology |
| STING (E9X7F) Rabbit mAb | #90947 | Cell Signaling Technology |
| Phospho-STING (Ser366) (E9A9K) Rabbit mAb | #50907 | Cell Signaling Technology |
| cGAS (E5V3W) Rabbit mAb | #79978 | Cell Signaling Technology |
| Phospho-Histone H2A.X (Ser139) (D7T2V) Mouse mAb | #80312 | Cell Signaling Technology |
| Anti-rabbit IgG (H+L), F(ab')2 Fragment (Alexa Fluor® 488 Conjugate) | #4412 | Cell Signaling Technology |
| Anti-mouse IgG (H+L), F(ab')2 Fragment (Alexa Fluor® 594 Conjugate) | #8890 | Cell Signaling Technology |
| ds DNA Marker Antibody (HYB331-01) | sc-58749 | Santa Cruz Biotechnology |

**Table S4:** Antibodies used for immunohistochemistry staining of patient derived FFPE tissue slides from the MUI cohort.

| **Target** | **Antibody** | Company |
|---|---|---|
| γH2AX (DNA-damage) | AF2288 | R&D Systems |
| STING (cGAS_STING activation) | HPA038116 | Sigma Aldrich |
| CD8 (CD8+ Tcells) | HPA037756 | Sigma Aldrich |
| PD-1 (Exhaustion) | HPA035981 | Sigma Aldrich |
| M2 macrophages CD163 | HPA046404 | Sigma Aldrich |
| CD4 (CD4+ T cells) | HPA004472 | Sigma Aldrich |
| FOXP3 (regulatory T-cells) | 98377 | CellSignaling |

**Table S5:** Immune related gene signatures.

| Signature | Ref. | Genes |
|---|---|---|
| T-cell exclusion | (1) | *BZW2, CCT3, CDK4, GPATCH4, ISYNA1, MDH2, PPIA, RPL31, RPL37A, RPL41, RPS21, RPS27A, RUVBL2, SAE1, UBA52, AHCY, C19orf48, C1QBP, CCT6A, CHCHD2, CTPS1, EEF1G, EIF2S3, EIF3K, EIF4A1, FARSA, FBL, FKBP4, GAS5, GNB2L1, GPI, HNRNPA1, HNRNPC, IDH2, ILF2, NACA, NCL, NME1, NOLC1, PABPC1, PAICS, PFN1, POLD2, PPA1, PTMA, PUF60, RPL10A RPL11, RPL12, RPL13, RPL13A, RPL13AP5, RPL17, RPL18, RPL18A, RPL21, RPL26, RPL27, RPL28, RPL29, RPL3, RPL36, RPL36A, RPL37, RPL4, RPL5, RPL6, RPL8, RPLP0, RPLP1, RPS10, RPS11, RPS13, RPS14, RPS15, RPS15A RPS16, RPS17, RPS17L, RPS18, RPS19, RPS23, RPS24, RPS27, RPS28, RPS3, RPS4X, RPS5, RPS6, RPS7, RPS8, RPS9, RPSA, RRS1, SERPINF1, SET, SHMT2, SLC19A1, SLC25A13, SNHG6, SNRPE, SOX4, SSR2, TIMM50, TOP1MT, TUBB, UQCRFS1, UQCRH, VDAC2, APP, ATP5D, ATP5G3, BOP1, BTF3, C6orf48, CACYBP, CCT4, CCT7, CDCA7, DARS, DCTPP1, DDX21, EEF1B2, EEF1D, EEF2, EIF3E, EIF3F, EIF3G, EIF3M, ENO1, EXOSC5, FAM92A1, GGH, GNL3, HMGB1, HNRNPH1, HNRNPM, HSPD1, IFRD2, ILF3, IMPDH2, LDHB, LSM4, LSM7, LYPLA1, MAGEC1, MCM7, MKI67IP, MRPL15, MRPL37, MRPL4, MRPS12, NDUFA11, NME2, NOP16, NPM1, NREP, PLEKHJ1 POLR1D, POLR2E, PRMT1, RPL10, RPL14, RPL15, RPL19, RPL22, RPL27A, RPL30, RPL32, RPL35, RPL39, RPL7, RPL7A, RPL9, RPLP2, RPS2, RPS20, RPS25, RPS3A, RQCD1, RSL1D1, SERBP1, SLC25A6, SMARCA4, SMIM15, SNHG15, SNRPB, SNRPC, SNRPD1, SNRPD2, SRM, SSB, TIMM13, TIMM44, TPI1, TRAP1, TRIM28, TYMS, UCK2, UHRF1, XIST, ZFAS1* |
| Inflamed | (2) | *IRF1, CD8A, CCL2, CCL3, CCL4, CXCL9, CXCL10, ICOS, GZMK, HLA-DMA, HLA-DMB, HLA-DOA, HLA-DOB* |
| Expand immune | (3) | *CXCR6, CD3D, CD2, ITGAL, TAGAP, CIITA, HLA-DRA, PTRPC, CXCL9, CCL5, NKG7, GZMA, PRF1, CCR5, CD3E, GZMK, IFNG, HLA-E, GZMB, PDCD1, SLAMF6, CXCL13, CXCL10, IDO1, LAG3, STAT1, CXCL11* |
| IFNG | (3) | *IFNG, STAT1, CCR5, CXCL9, CXCL10, CXCL11, IDO1, PRF1, GZMA, HLA-DRA* |
| CTL | (4) | *CD8A, CD8B, GZMA, GZMB, PRF1* |
| CD8 | (4) | *CD8A, CD8B* |
| CYT | (5) | *GZMA, PRF1* |
| CD8 T-cell exhaustion | (6) | *PDCD1, LAYN, HAVCR2, LAG3, CD244, CTLA4, LILRB1, TIGIT, TOX, VSIR, BTLA, ENTPD1, CD160, LAIR1* |
| Core NFkB | MSigDB: M8804 (7) | *BCL3, CHUK, NFKBIA, NFKBIB, NFKBIE, IKBKB, IKBKE, IKBKG, NFKB1, NFKB2, REL, RELA, RELB* |
| IRF3 targets | MSigDB: M5133 (8) | *B4GALT5, IFIT3, ISG15, IFI44, ARG2, PMAIP1, GBP1, F13B, AHNAK, OAS2, NR3C1, IFIT1, PLCG2* |
| STING signaling | MSigDB: M7982 (9) | *ADAR, IFNA8, IFNA1, IFNB1, IRF1, IRF2, IRF6, IRF7, IRF9, CCL5* |
| Interferon α response | MSigDB: M5911 (10) | *PARP12, PARP14, PARP9, PLSCR1, PNPT1, PROCR, PSMA3, PSMB8, PSMB9, PSME1, PSME2, RIPK2, RNF31, RSAD2, RTP4, SAMD9, SAMD9L, SELL, SLC25A28, SP110, STAT2, TAP1, TDRD7, TENT5A, TMEM140, TRAFD1, TRIM14, TRIM21, TRIM25, TRIM26, TRIM5, TXNIP, UBA7, UBE2L6, USP18, WARS1* |
| Integrins | (11) | *ITGAD, ITGAE, ITGAL, ITGAM, ITGAV, ITGAX, ITGA1, ITGA2, ITGA2B, ITGA3, ITGA4, ITGA5, ITGA6, ITGA7, ITGA8, ITGA9, ITGA10, ITGA11, ITGBL1, ITGB1, ITGB2, ITGB3, ITGB4, ITGB5, ITGB6, ITGB7, ITGB8* |
| Chemokines | (12) | *CCL1, CCL2, CCL3, CCL3L1, CCL3L3, CCL4, CCL4L1, CCL4L2, CCL5, CCL7, CCL8, CCL11, CCL13, CCL14, CCL15, CCL16, CCL17, CCL18, CCL19, CCL20, CCL21, CCL22, CCL23, CCL24, CCL25, CCL26, CCL27, CCL28, CXCL1, CXCL2, CXCL3, CXCL5, CXCL6, CXCL8, CXCL9, CXCL10, CXCL11, CXCL12, CXCL13, CXCL14, CXCL16, CXCL17, CX3CL1, XCL1, XCL2* |
| Cytokines | (13) | *IL1A, IL1B, IL1F10, IL1RN, IL2, IL3, IL4, IL5, IL6, IL7, CXCL8, IL9, IL10, IL11, IL12A, IL12B, IL13, IL15, IL16, IL17A, IL17B, IL17C, IL17D, IL17F, IL18, IL19, IL20, IL21, IL22, IL23A, IL24, IL25, IL26, IL27, IL31, IL32, IL33, IL34, IL36A, IL36B, IL36G, IL36RN, IL37* |
| HLAs | (14) | *HLA-A, HLA-B, HLA-C, HLA-DMA, HLA-DMB, HLA-DOA, HLA-DOB, HLA-DPA1, HLA-DPB1, HLA-DQA1, HLA-DQA2, HLA-DQB1, HLA-DQB2, HLA-DQB3, HLA-DRA, HLA-DRB1, HLA-DRB3, HLA-DRB4, HLA-DRB5, HLA-E, HLA-F, HLA-G* |

**Table S6:** Lists of selected genes associated with different immune related functions corresponding to Figure 4A.

| Signature | Genes |
|---|---|
| Cytotoxic effector function | *CD8A, CD8B, EOMES, FASLG, GNLY, GZMA, GZMB, GZMK, IFNG, PRF1, TBX21, ZAP70* |
| Immune response | *IRF1, CD8A, CCL2, CCL3, CCL4, CXCL9, CXCL10, ICOS, GZMK, HLA-DMA, HLA-DMB, HLA-DOA, HLA-DOB* |
| Antigen processing and presentation | *B2M, CD1C, CD1D, CD74, CD83, CIITA, HLA-A, HLA-B, HLA-C, HLA-DMA, HLA-DMB, HLA-DOA, HLA-DOB, HLA-DPA1, HLA-DPB1, HLA-DQA1, HLA-DQA2, HLA-DQB1, HLA-DQB2, HLA-DRA, HLA-DRB1, HLA-E, HLA-F, HLA-G, MR1, PSMB7, PSMB8, PSMB9, TAP1, TAP2, TAPBP* |
| Immune checkpoints | *ADORA2A, BTLA, BTNL2, CD160, CD2, CD200, CD200R1, CD226, CD244, CD27 CD274, CD276, CD28, CD40, CD40LG, CD47, CD48, CD58, CD69, CD70, CD80, CD86, CD96, CEACAM1, CTLA4, ENTPD1, EOMES, HAVCR1, HAVCR2, HHLA2, ICOS, ICOSLG, IDO1, IDO2, KDR, KIR2DL4, KLRC1, KLRK1, LAG3, LAIR1, LGALS9, LTA, MICA, MICB, NCR3, NCR3LG1, NECTIN2, NECTIN3, NT5E, PDCD PDCD1LG2, PVR, RAET1E, SIRPA, SIVA1, SLAMF1, SLAMF6, TDO2, TIGIT, TIMD4, TMEM173, TMIGD2, TNFRSF13B, TNFRSF13C, TNFRSF14, TNFRSF17, TNFRSF18, TNFRSF25, TNFRSF4, TNFRSF8, TNFRSF9, TNFSF13, TNFSF13B, TNFSF14, TNFSF15, TNFSF18, TNFSF4, TNFSF8, TNFSF9, TRAF1, TRAF2, TRAF3, ULBP1, VSIR, VTCN1* |
| INFG | *STAT1, CCR5, CXCL9, CXCL10, CXCL11, IDO1, PRF1, GZMA, HLA-DRA* |
| T-cell exhaustion | *HAVCR2, PDCD1, ENTPD1, TNFRSF9, SIRPG, CTLA4, CXCL13, TOX, MYO7A, VCAM1, TIGIT, LAG3, NDFIP2, ACP5, DUSP4, MIR155HG, PHLDA1, CCL3, IFNG, GZMB, PARK7, CXCR6, CD27, FKBP1A, BST2, CD63, CD27-AS1, ITGAE, HLA-DRA, IGFLR1, CTSD, CD38, ITM2A, SNAP47, LAYN, KRT86, TNFRSF18, RBPJ, CD82, RGS1* |
| TGFb/CAF marker | *RCN3, ZCCHC24, BMP4, TNFRSF8, TDO2, INHBA, NTM, FAP, ACTA2, ASPN, COL11A1, CSPG4, DES, FAP, FN1, FOXF1, ITGA11, ITGB1, MFAP5, MME, OGN, P4HA1, P4HB, PDGFRA, PDGFRB, PDPN, POSTN, S100A4, SLC16A4, SPARC, THY1, TNC, VIM, ZEB1* |

**Table S7:** Results of multivariable survival analysis using Cox regression for BRCAness in the TCGA cohort.

| | HR | 95% confidence | p | |
|---|---|---|---|---|
| BRCAness | 0.488 | (1.40-2.99) | 0.00023 | *** |
| Tumor residual disease | 2.919 | (1.55-5.48) | 0.00086 | *** |
| Age | 1.027 | (1.01-1.05) | 0.0024 | ** |
| FIGO Stage 2 vs.3 | 0.956 | (0.30-3.08) | 0.9401 | |
| FIGO Stage 3 vs 4 | 1.426 | (0.42-4.83) | 0.5682 | |

**Table S8:** Results of multivariable survival analysis using Cox regression for the vulnerability score in the TCGA cohort.

| | HR | 95% confidence | p | |
|---|---|---|---|---|
| Vulnerability score | 0.754 | (0.65-0.87) | 0.00015 | *** |
| Tumor residual disease | 2.963 | (1.58-5.56) | 0.00071 | *** |
| Age | 1.027 | (1.01-1.04) | 0.0023 | ** |
| FIGO Stage 2 vs.3 | 0.964 | (0.30-3.10) | 0.9516 | |
| FIGO Stage 3 vs 4 | 1.446 | (0.43-4.90) | 0.5529 | |

**Table S9:** Results of multivariable survival analysis using Cox regression for CD8 T-cell infiltration in the TCGA cohort.

| | HR | 95% confidence | p | |
|---|---|---|---|---|
| CD8+ T-cell infiltration | 0.00043 | (0.00-1.95E+06) | 0.494 | |
| Tumor residual disease | 3.286 | (1.75-6.16) | 0.00021 | *** |
| Age | 1.027 | (1.01-1.05) | 0.0022 | ** |
| FIGO Stage 2 vs.3 | 1.129 | (0.35-3.60) | 0.8379 | |
| FIGO Stage 3 vs 4 | 1.515 | (0.45-5.08) | 0.5015 | |

### Supplementary Figures

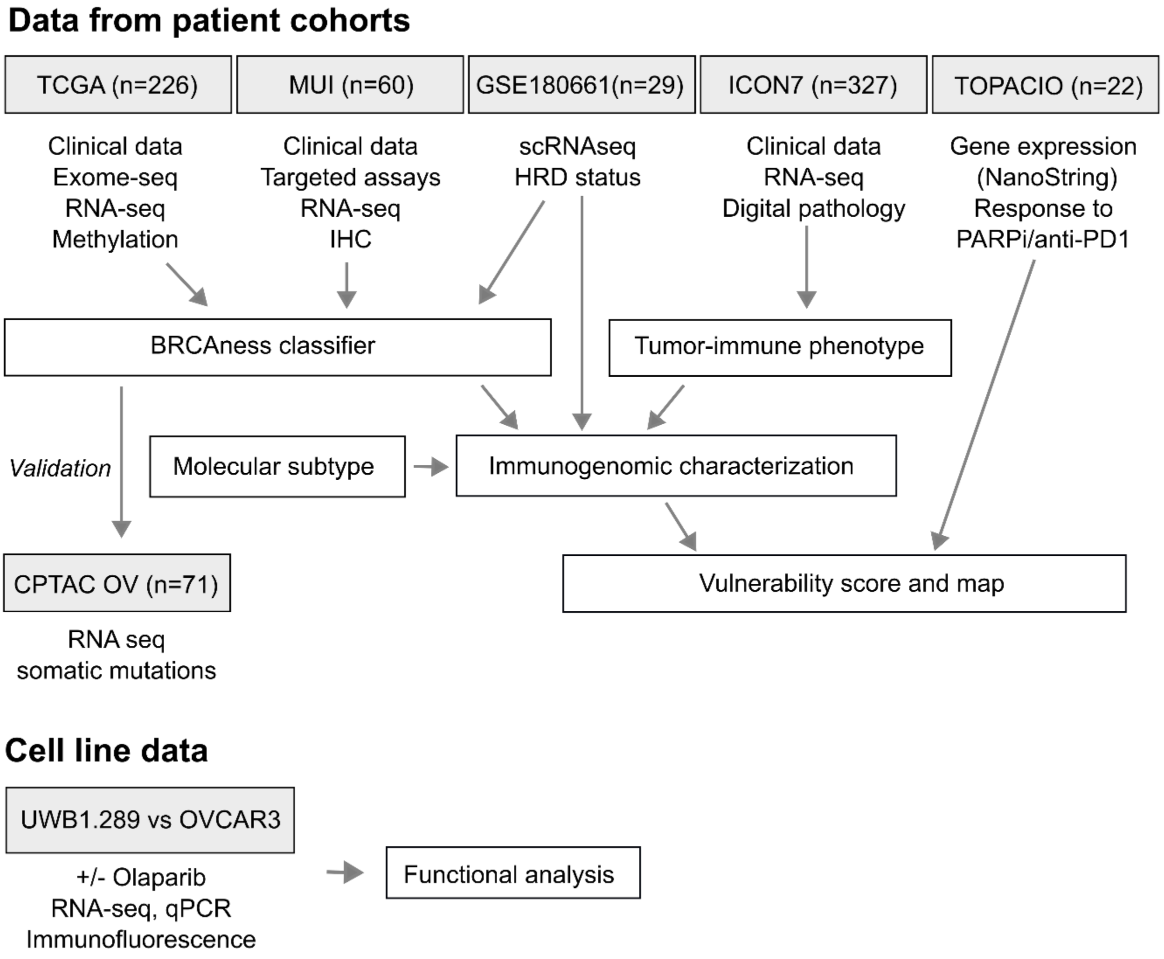


**Figure S1:** Workflow of analyses and ovarian cancer patient cohorts with corresponding data sets used for immunogenomic characterization of patient samples and ovarian cancer cell line data. IHC, immunohistochemistry; OV, ovarian cancer; HRD, homologous recombination repair deficiency; PARPi, PARP inhibitor; qPCR, quantitative reverse transcription polymerase chain reaction; scRNAseq, single cell RNA sequencing; CPATC, Clinical Proteomic Tumor Analysis Consortium; TCGA, The Cancer Genome Atlas; MUI, Medical University of Innsbruck; GSE, Gene Expression Omnibus Identifier.

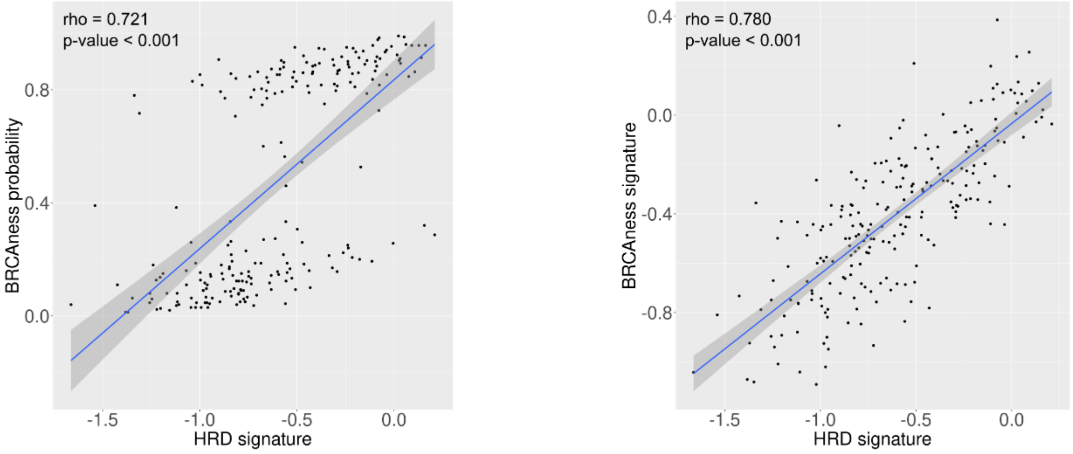


**Figure S2:** Correlation of different BRCAness and HRD-parameter in the TCGA cohort. BRCAness prediction probability and HRD signature (15) (left), BRCAness signature and HRD signature (15) (right).

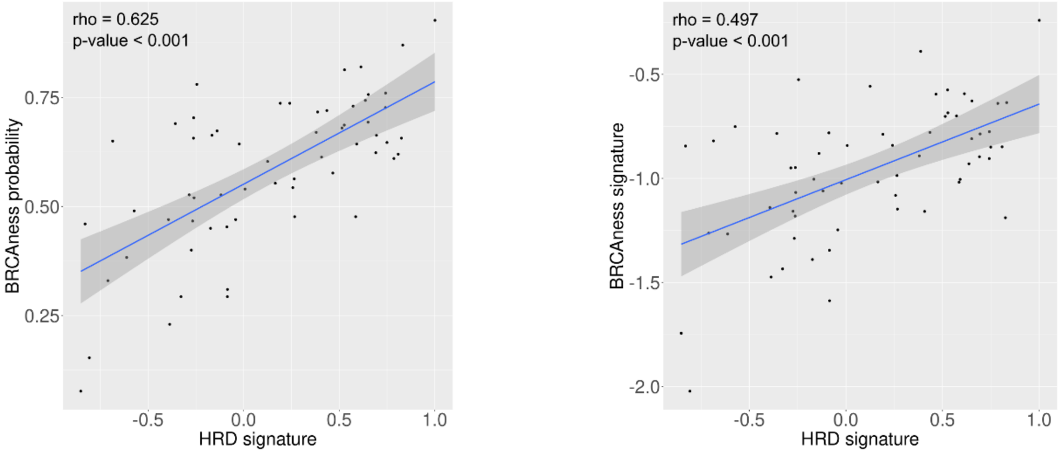


**Figure S3:** Correlation of different BRCAness and HRD-parameter in the MUI cohort. (left) BRCAness prediction probability and HRD signature (15), BRCAness signature and and HRD signature (15).

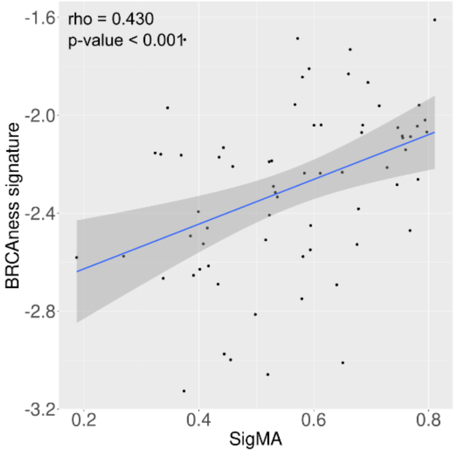


**Figure S4:** Correlation of the BRCAness signature and SigMA (Mutational signature 3) in the CPTAC-OV cohort.

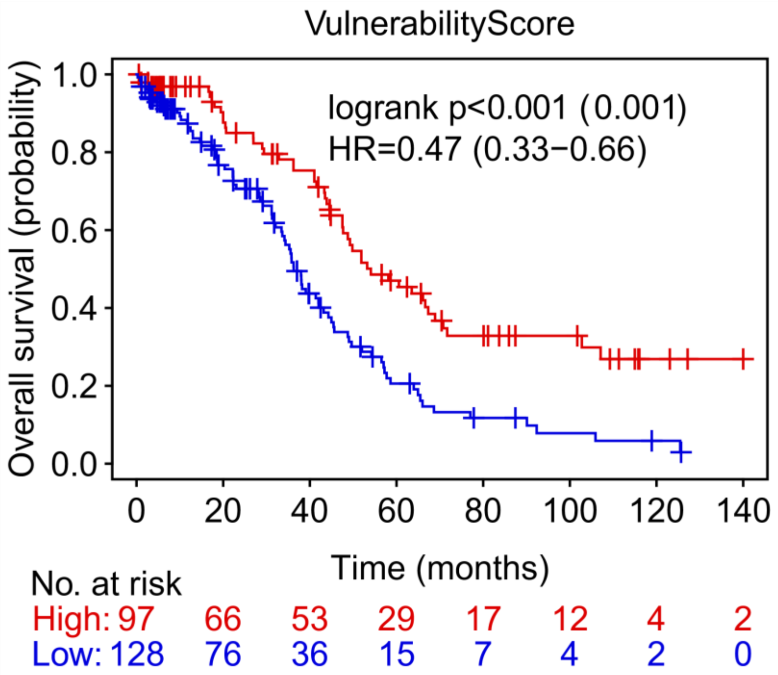


**Figure S5:** Kaplan Meier curve for the vulnerability score in the TCGA cohort. Patients were dichotomized using maximum logrank statistics/minimal p-value. Corrected p-values were calculated according to Altman et al. (16) and indicated in brackets. TCGA, The Cancer Genome Atlas.

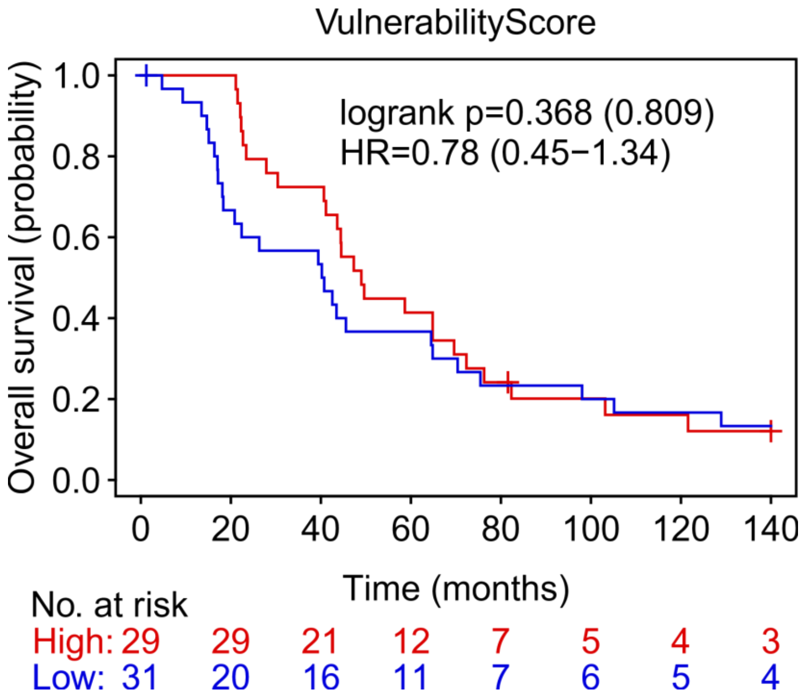


**Figure S6:** Kaplan Meier curve for the vulnerability score in the MUI cohort. Patients were dichotomized using maximum logrank statistics/minimal p. Corrected p-values were calculated according to Altman et al. (16) and indicated in brackets. MUI, Medical University of Innsbruck.

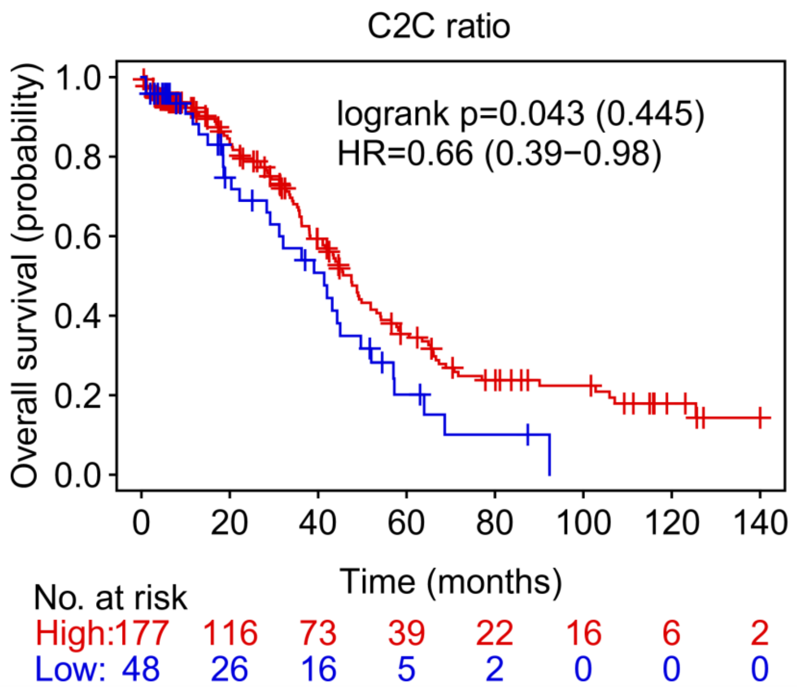


**Figure S7:** Kaplan Meier curve for the CYT to *C1QA* ratio (C2C) in the TCGA cohort. Patients were dichotomized using maximum logrank statistics / minimal p-value. Corrected p-values were calculated according to Altman et al. (16) and indicated in brackets. TCGA, The Cancer Genome Atlas.

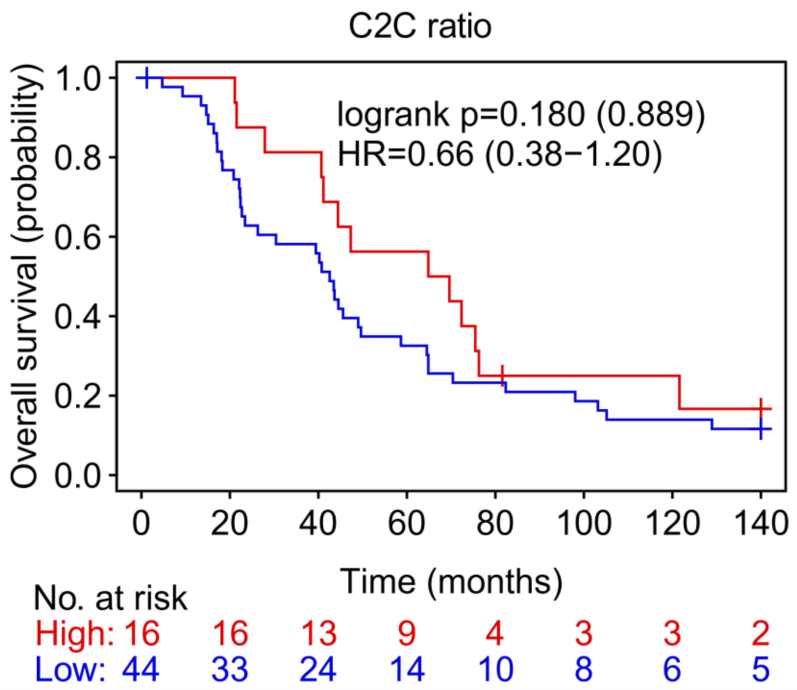


**Figure S8:** Kaplan Meier curve for the CYT to *C1QA* ratio (C2C) in the MUI cohort. Patients were dichotomized using maximum logrank statistics / minimal p-value. Corrected p-values were calculated according to Altman et al. (16) and indicated in brackets. MUI, Medical University of Innsbruck.

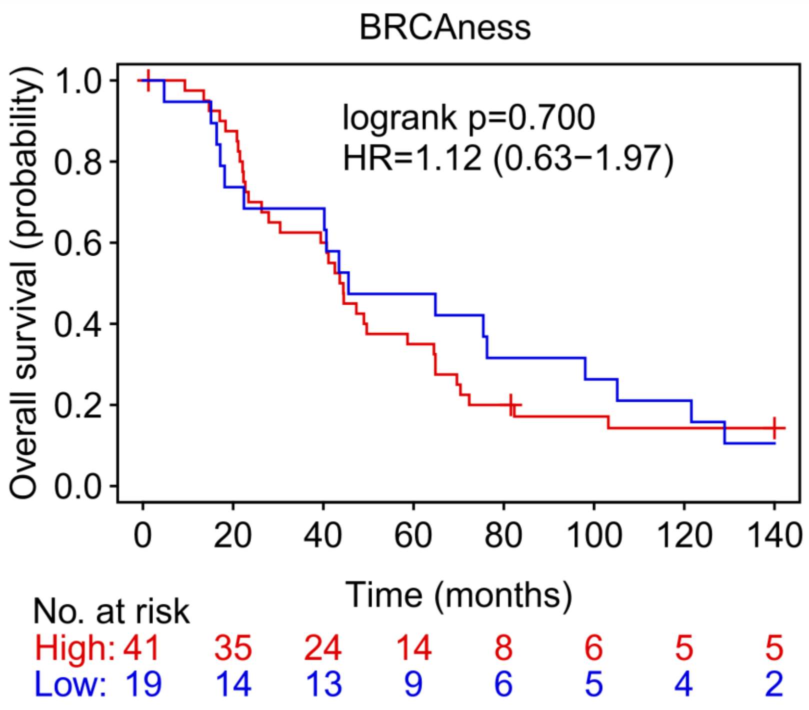


**Figure S9:** Kaplan Meier curve for BRCAness (red) versus noBRCAness (blue) in the MUI cohort. MUI, Medical University of Innsbruck.

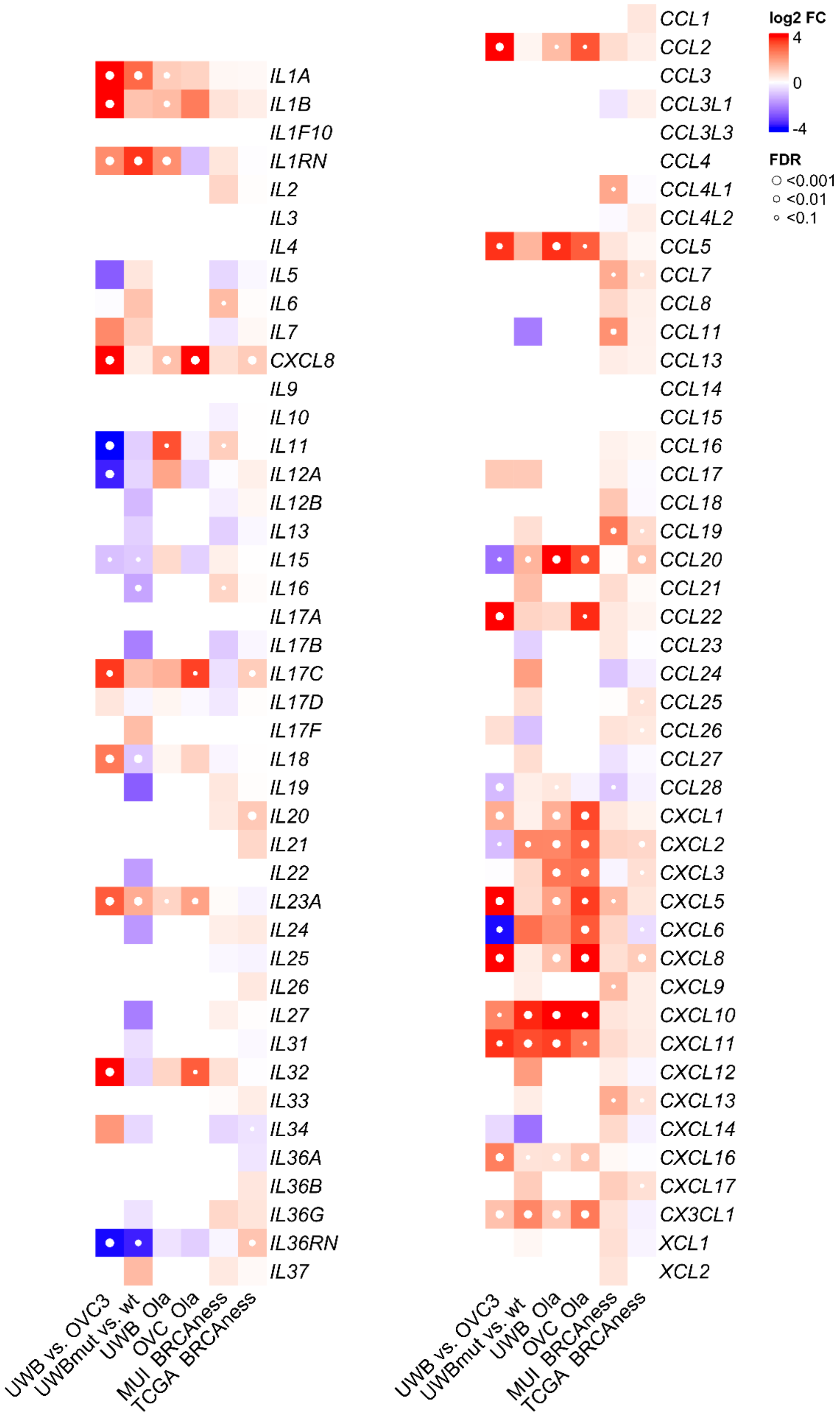


**Figure S10:** Differential expressed cytokines and chemokines between BRCAness noBRCAness patients as well as by olaparib treatment in cell lines. UWB, cell line UWB1.289; OVC, cell line OVCAR3; TCGA, The Cancer Genome Atlas; MUI, Medical University of Innsbruck; Ola, Olaparib treatment; wt, wilde type; mut, BRCA1 mutation.

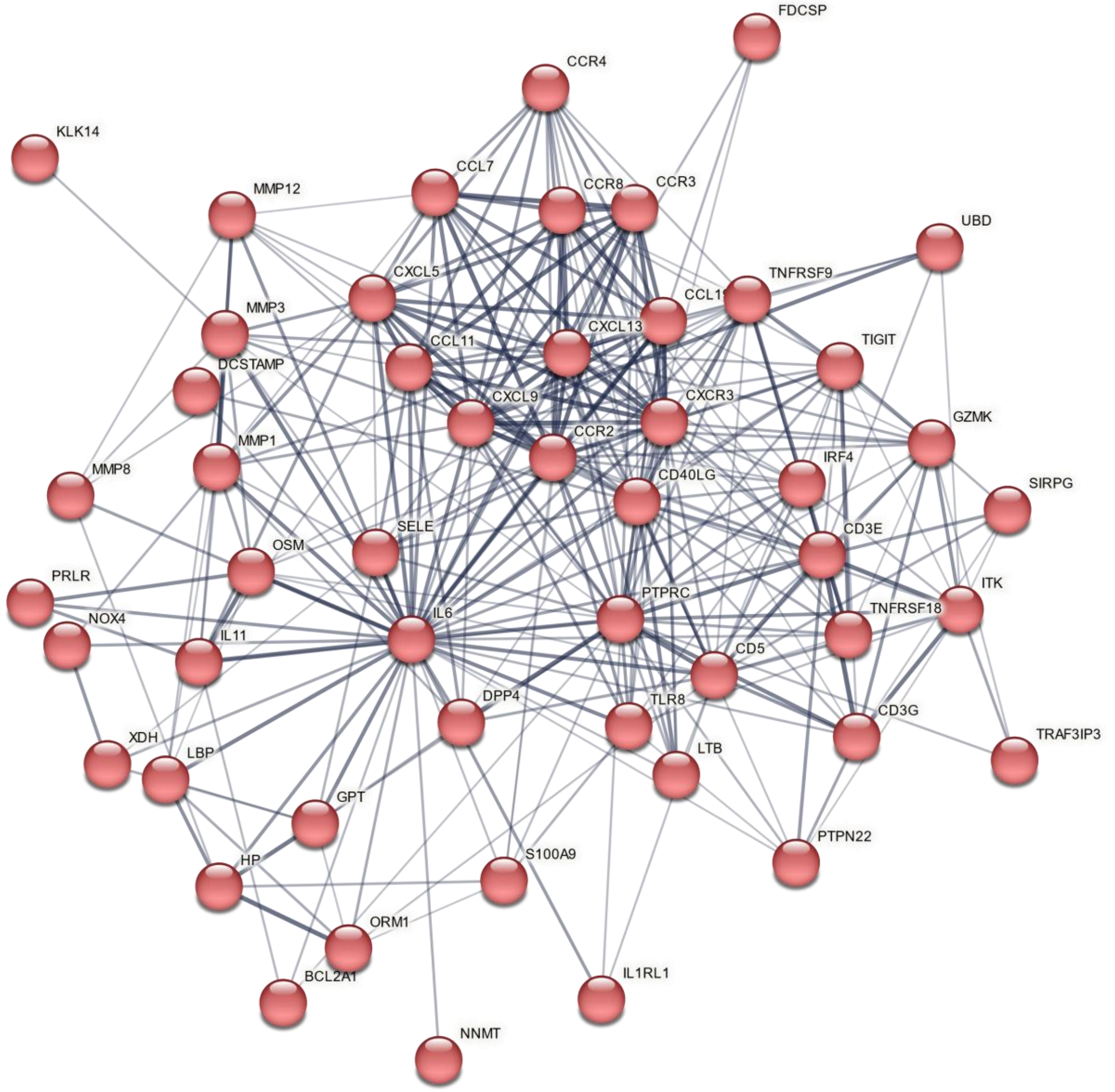


**Figure S11:** Network cluster of genes associated with immune signaling pathways which are upregulated in the BRCAness samples of the MUI Cohort. MUI, Medical University of Innsbruck.

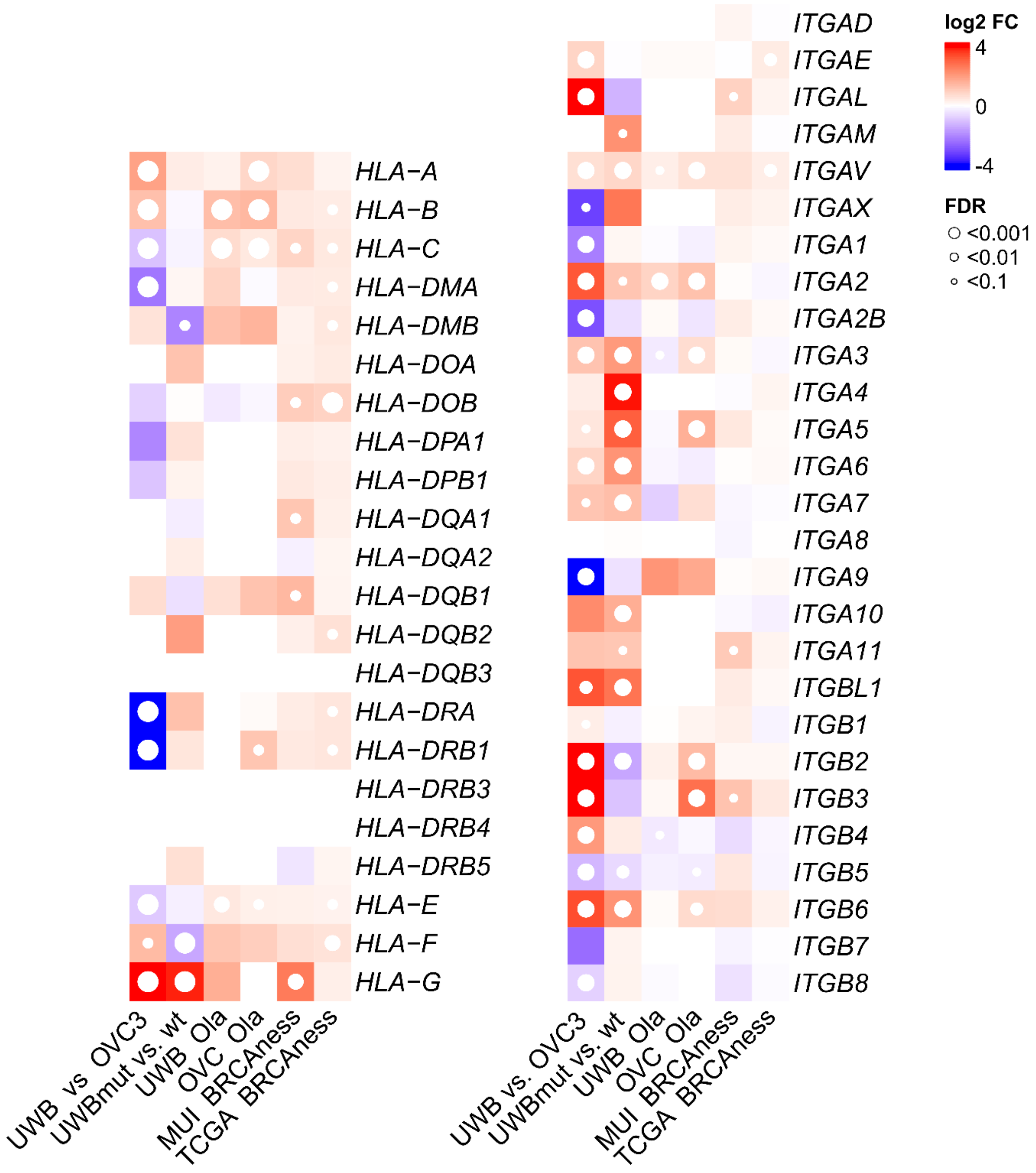


**Figure S12:** Differentially expressed HLA-genes and integrins between BRCAness noBRCAness patients as well as by olaparib treatment in cell lines. UWB, cell line UWB1.289; OVC, cell line OVCAR3; TCGA, The Cancer Genome Atlas; MUI, Medical University of Innsbruck; Ola, Olaparib treatment; wt, wilde type; mut, BRCA1 mutation.

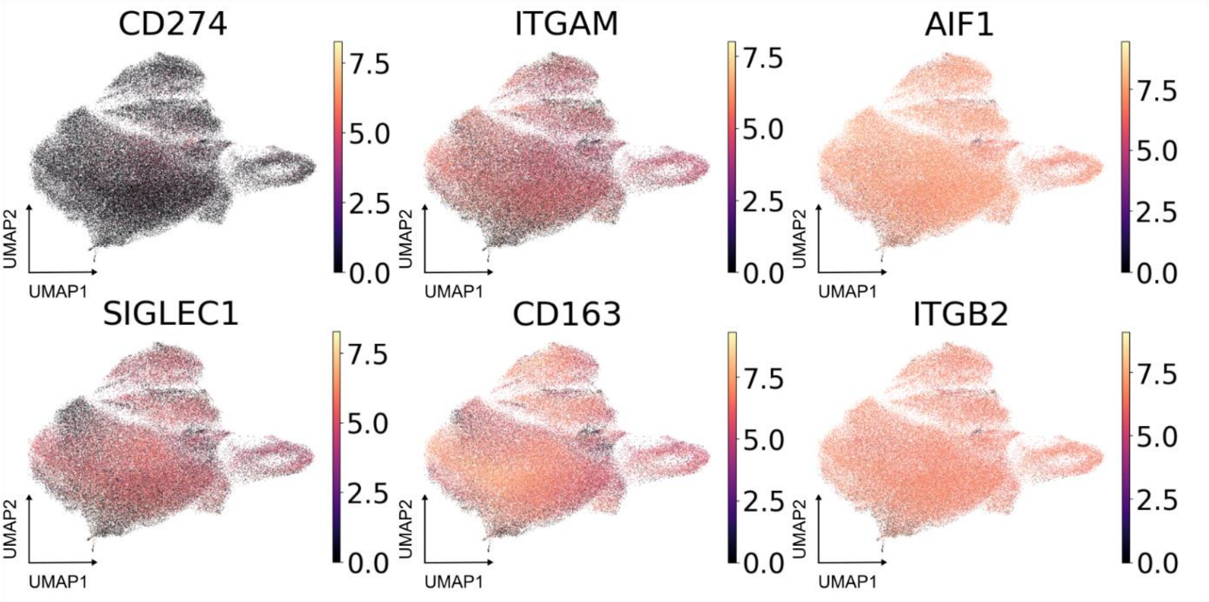


**Figure S13:** UMAP plots of selected macrophage associated genes in the myeloid cell compartment of single cell RNAseq analyses.

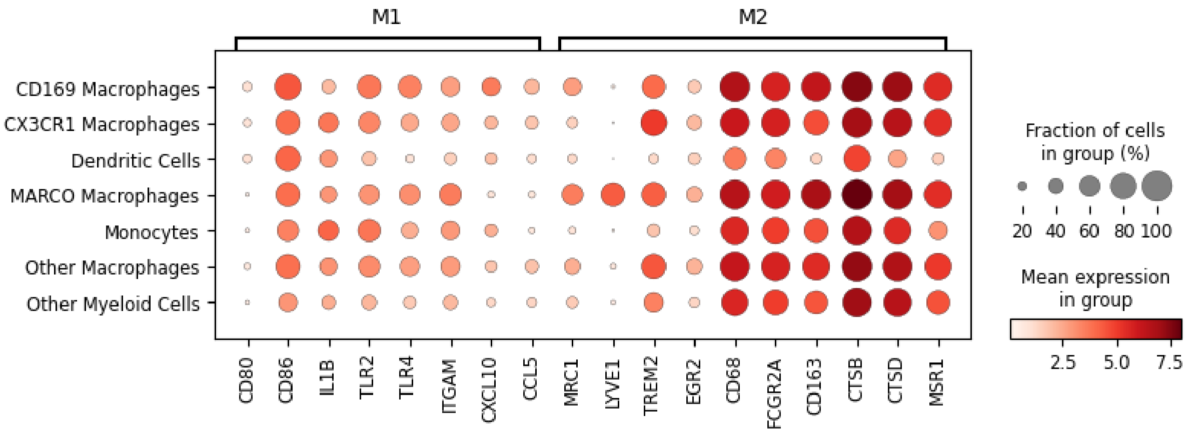


**Figure S14:** Expression of marker genes for macrophage polarization in the myeloid subgroups of the ovarian cancer microenvironment. Expression of M1 marker genes (left) and M2 marker genes right (M2) indicate a bias towards M2 macrophages, although discrimination is limited.

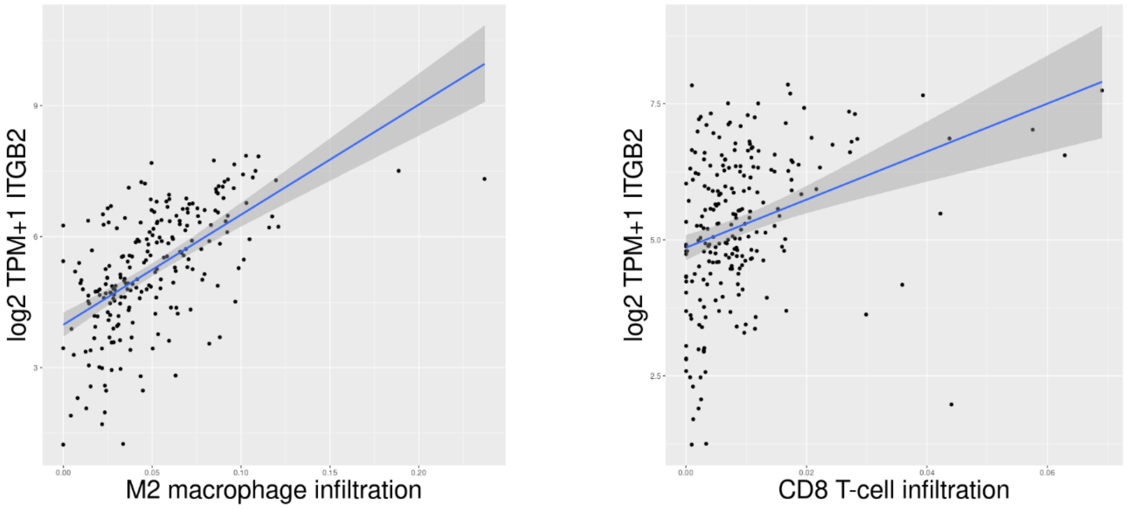


**Figure S15:** Correlation of M2 macrophage (left) and CD8 T-cell infiltration (right) and *ITGB2* expression in the TCGA cohort. TCGA, The Cancer Genome Atlas.

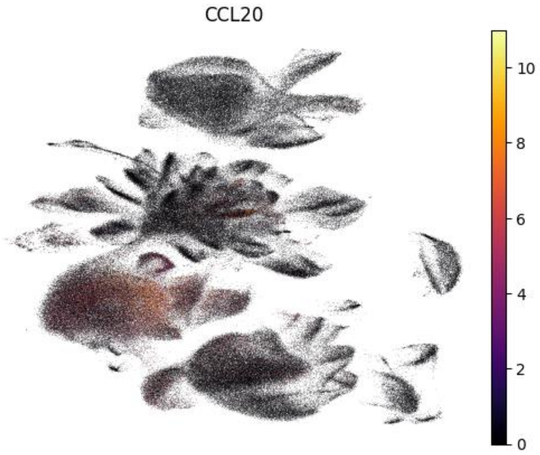


**Figure S16:** Expression of *CCL20* in myeloid cells and to a less extent in some tumor cells of BRCAness samples.

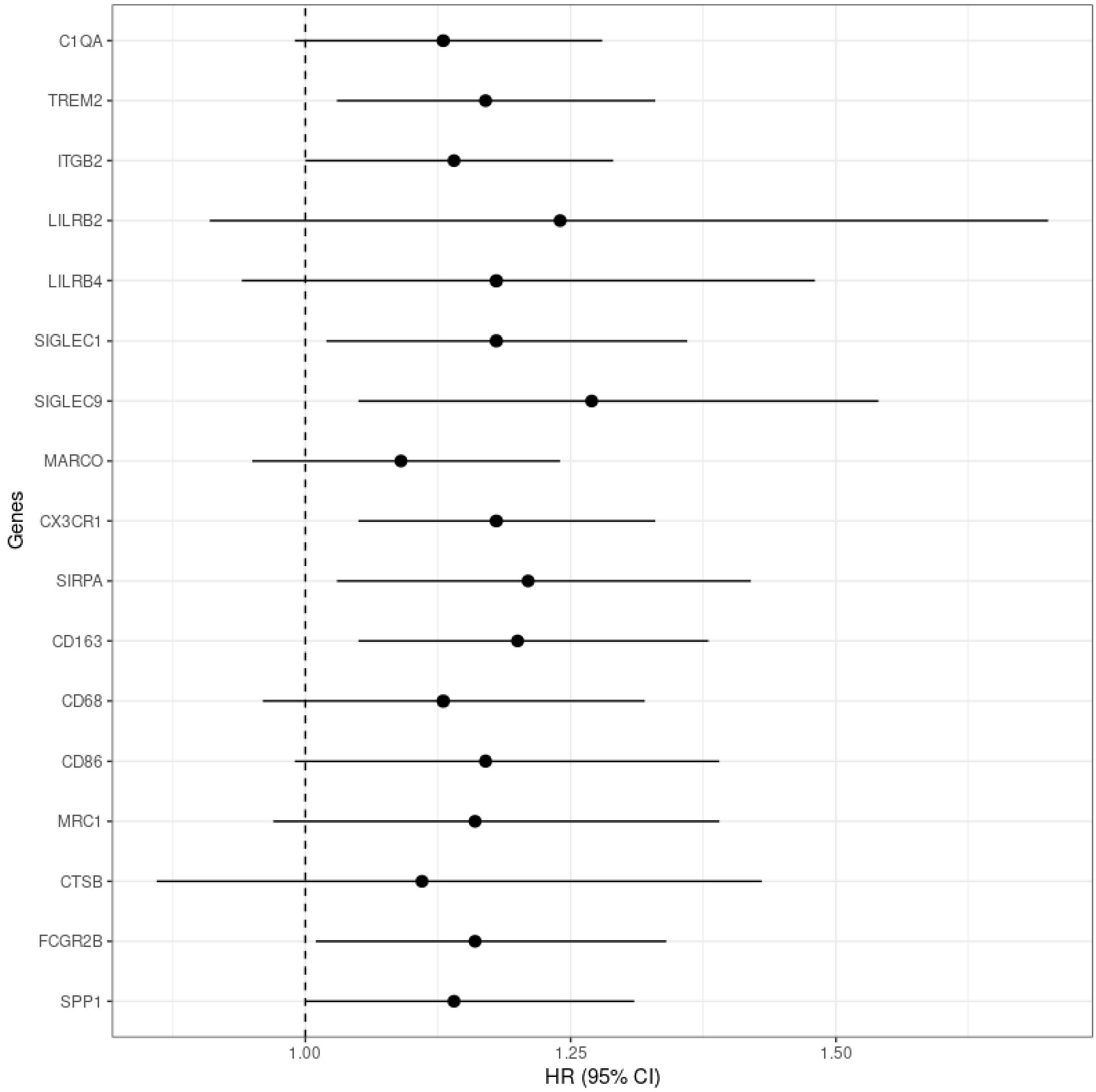


**Figure S17:** Forest plot showing the hazard ratio and the 95% CI for selected genes associated with tumor associated macrophages in the TCGA cohort using Cox regression and including BRCAness status as covariate. CI, confidence interval.

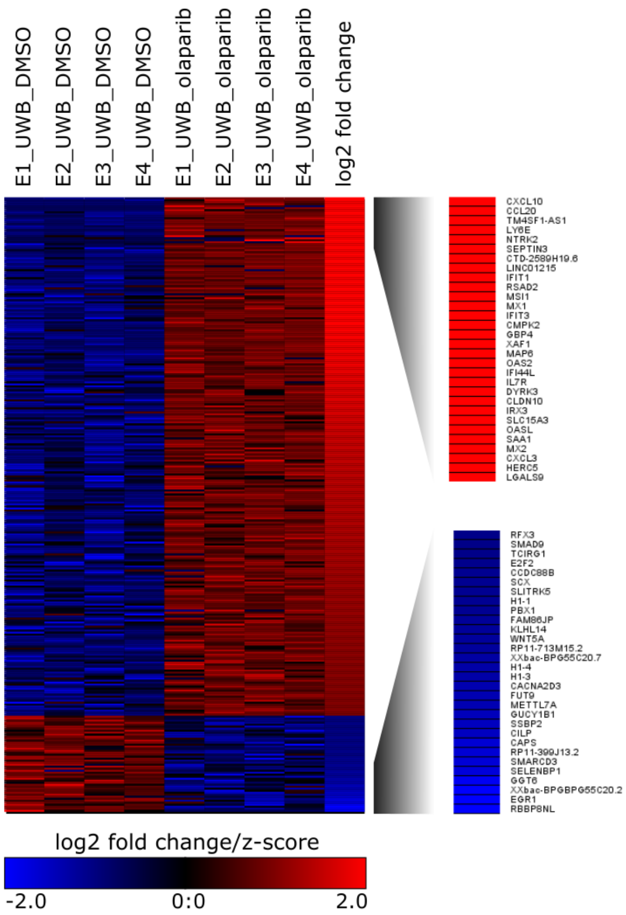


**Figure S18:** Z-scores of normalized counts and log2-fold changes of significant differential expressed genes between olaparib treated and control treated samples of the UWB1.289 cell line.

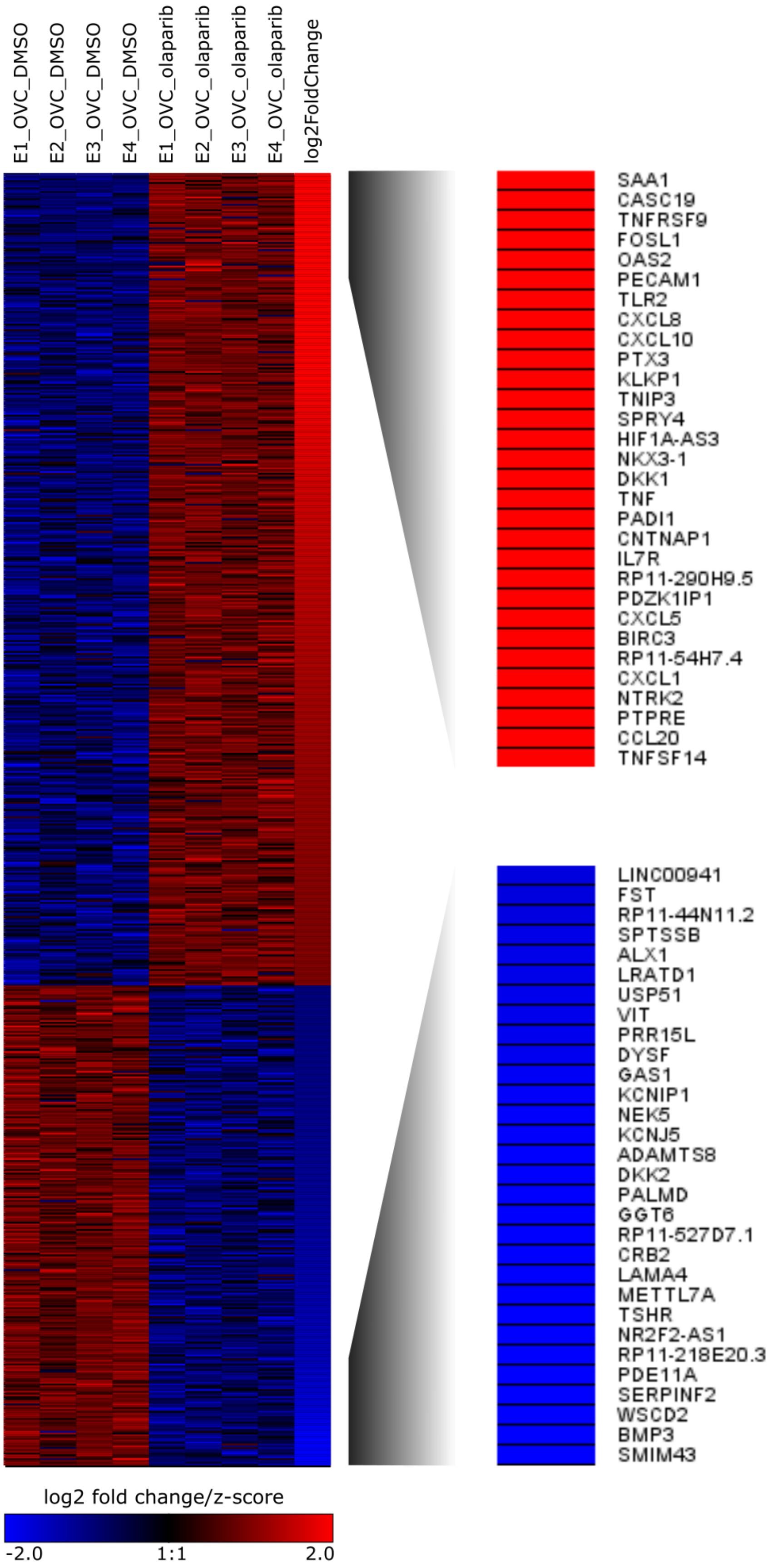


**Figure S19:** Heatmap of z-scores of normalized counts and log2-fold changes of significant differential expressed genes between olaparib treated and control treated samples of the OVCAR3 cell line.

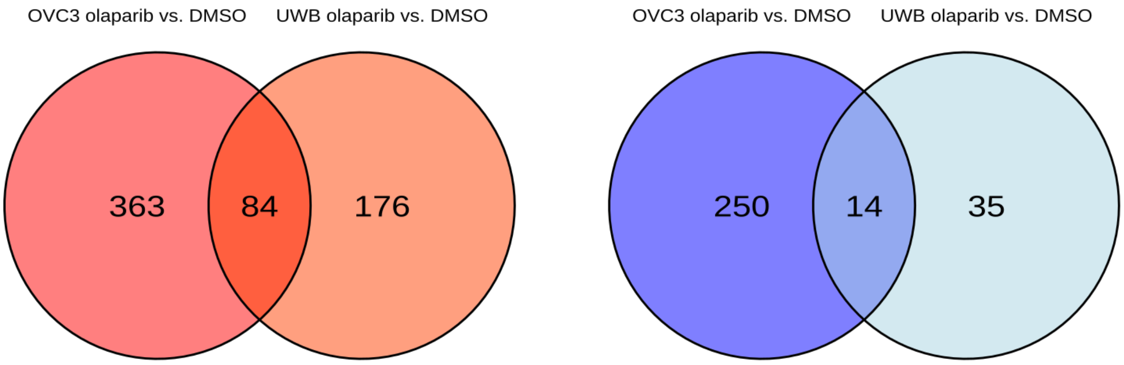


**Figure S20:** Number and overlap of significant up- and down regulated genes by olaparib treatment in the OVCAR3 and UWB1.289 ovarian cancer cell lines.

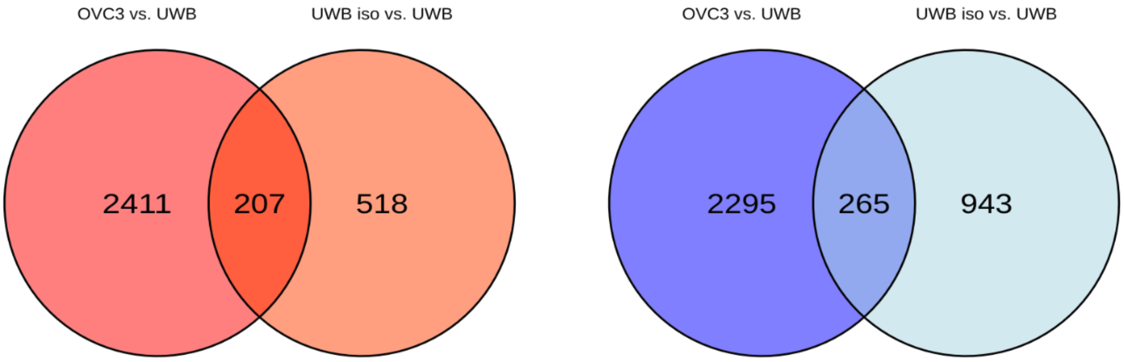


**Figure S21:** Number and overlap of significant differentially regulated genes between the UWB1.289 and OVCAR3 ovarian cancer cell lines respectively UWB1.289 and isogenic UWB with reintroduced wild type *BRCA1* gene.

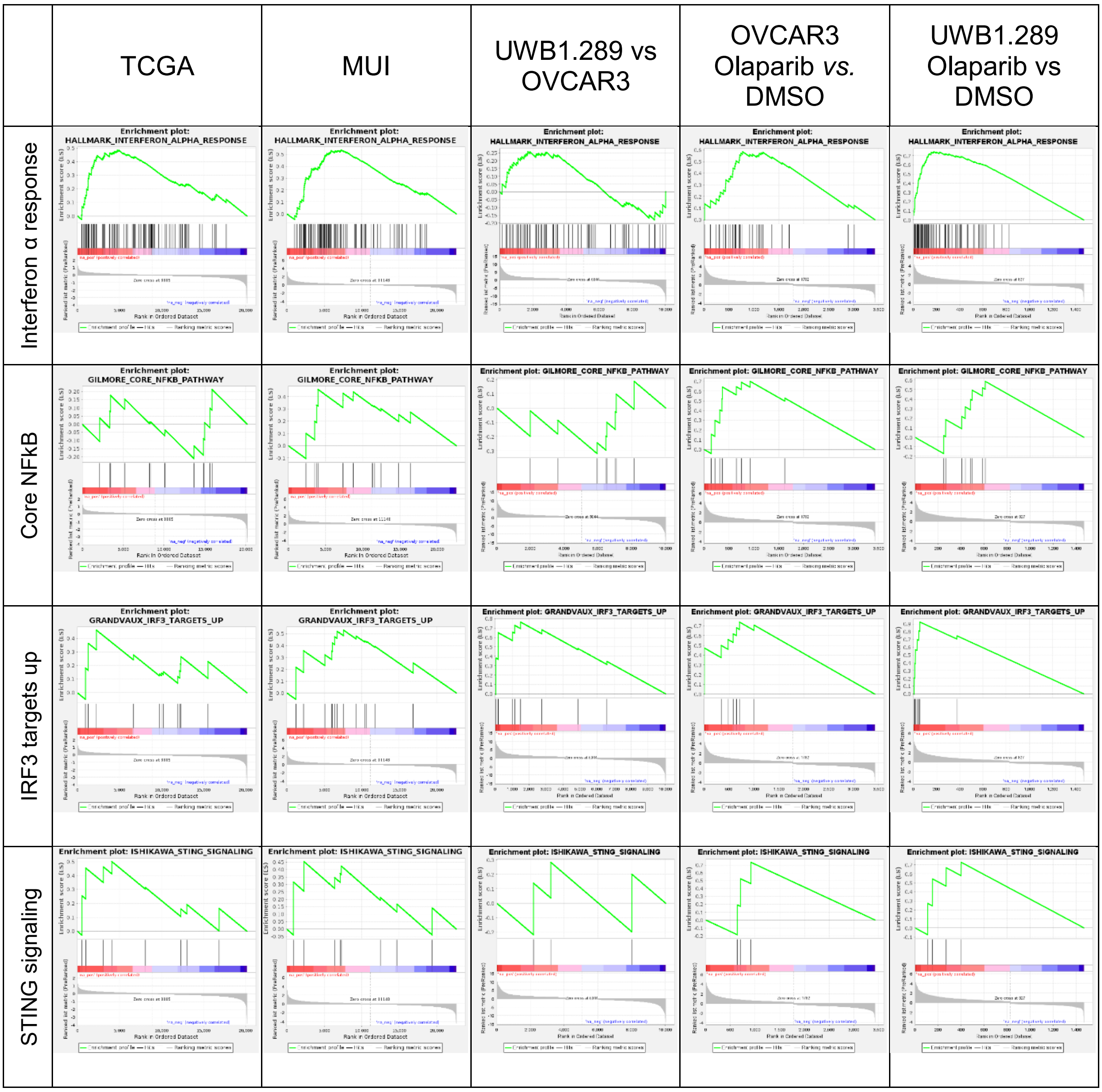


**Figure S22:** Enrichment plots for the pathways shown in Figure 3C, which are associated with the activation of the cGAS STING pathway.

**OvRSeq Analysis Report for MUI40**

2023-12-04

The vulnerability map indicate based on BRCAness probability and CYT to C1QA ratio (C2C) indications with a high vulnerability (score) for response to combination immunotherapy with PARPi and immune checkpoint inhibitors.

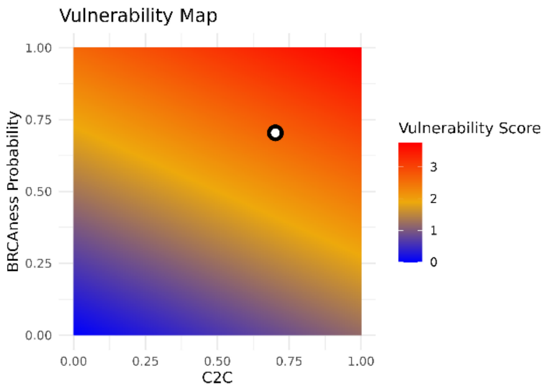


**Patient values**

| Feature | Value |
|---|---|
| BRCAness status | 1 (0.70) |
| Tumor immune phenotype | Infiltrated |
| Molecular subtypes | IMR |
| BRCAness immunetype | BRIT |
| Vulnerability score | 2.22 |
| Immunophenoscore | 5 |
| CYT to C1QA ratio (C2C) | 0.70 |
| Angiogenesis Score | 1 |

Marker gene expression and reference values from TCGA-OV [median (Q1-Q3)].

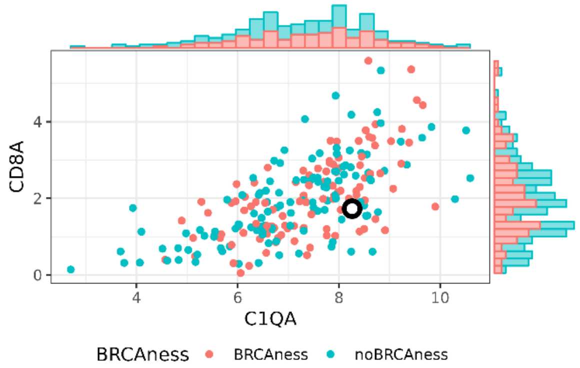


| Feature | Patient Value | TCGA IQR |
|---|---|---|
| CD274 | 1.91 | 1.32 (0.87-1.86) |
| GZMB | 3.30 | 2.55 (1.62-3.51) |
| PRF1 | 2.29 | 1.73 (1.15-2.50) |
| C1QA | 8.26 | 7.35 (6.41-8.18) |
| CD8A | 1.73 | 1.90 (1.17-2.65) |
| IDO1 | 4.52 | 4.16 (2.80-5.45) |
| FOXP3 | 1.59 | 1.94 (1.32-2.52) |
| TREM2 | 6.24 | 4.83 (3.76-5.64) |
| STAT1 | 7.24 | 7.01 (6.21-7.55) |
| HLA-DRA | 7.13 | 7.26 (6.30-8.00) |
| CXCL10 | 6.92 | 6.38 (5.03-7.54) |

### Enrichment of gene signatures or pathways (ssGSEA) and estimated immune cell fraction (quanTIseq)

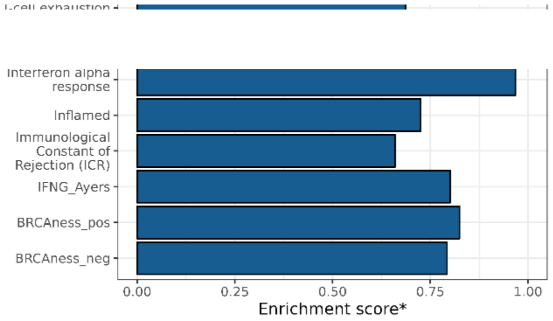


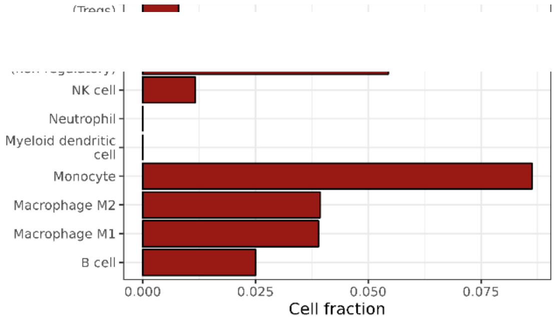


*Enrichment score is standardized to min-max values from TCGA-OV reference.

Enrichment score*

Cell fraction

1

**Figure S23:** Report for comprehensive characterization including vulnerability map and score of one HGSOC patient sample from RNA sequencing data using the web application https://ovrseq.icbi.at or the R package OvRSeq (https://github.com/icbi-lab/OvRSeq).

**References (supplement)**
